# Multimodal Integration of Ambulatory ECG and Clinical Features for Sudden Cardiac Death and Pump Failure Death Prediction^⋆^

**DOI:** 10.64898/2026.04.21.26351421

**Authors:** Steven Swee, Irsyad Adam, Erika Yilin Zheng, Ethan Ji, Ding Wang, William Speier, Jeffrey Hsu, Kai-Wei Chang, Kalyanam Shivkumar, Peipei Ping

**Affiliations:** Medical Informatics HA, University of California Los Angeles, USA; Department of Physiology, University of California Los Angeles, USA; Department of Computer Science, University of California Los Angeles, USA; Department of Radiological Sciences, University of California Los Angeles, USA; Department of Medicine / Cardiology Division, University of California Los Angeles, USA; Department of Biomedical Informatics, the University of Arizona College of Medicine, USA

**Keywords:** Multiple Instance Learning, TCN, LLM, Feature Fusion

## Abstract

Ambulatory electrocardiograms (ECGs) provide continuous monitoring of cardiac electrical activity, but many machine learning approaches analyze these recordings without incorporating broader patient clinical context. We evaluated a multimodal framework integrating ambulatory ECG-derived representations with clinical information for four-year prediction of sudden cardiac death (SCD) and pump failure death (PFD). The analysis included 730 patients from the MUSIC cohort, comprising 577 patients with no cardiac death, 71 with SCD, and 82 with PFD. Ambulatory ECG recordings were segmented and encoded using a multiple instance learning–temporal convolutional neural network, while clinical variables were represented as LLM-generated text, deterministic text, or tabular features. Models were developed and evaluated using patient-level nested cross-validation with five outer and four inner folds, with preprocessing, model and fusion-architecture selection, calibration, and threshold selection restricted to training data. For SCD, ECG + deterministic text achieved the highest AUROC of 0.711 (95% CI, 0.643–0.773), while ECG + tabular clinical features achieved the highest PR-AUC of 0.248 (0.180–0.357). For PFD, the unimodal tabular model achieved the highest AUROC of 0.787 (0.732–0.839) and PR-AUC of 0.360 (0.275–0.470), while ECG + tabular features achieved an AUROC of 0.756 (0.697–0.811) and PR-AUC of 0.353 (0.261–0.461). ECG + LLM text improved upon LLM text alone but did not outperform the unimodal ECG model. Attribution and token-masking analyses provided complementary evidence that model explanations were partly robust to superficial input perturbations while remaining sensitive to model and masking conditions, and representation and patient-text permutation analyses showed that predictive value depended strongly on how clinical information was represented. These findings indicate that the benefit of multimodal integration is outcome- and representation-dependent, with deterministic and tabular clinical representations providing greater predictive value than full LLM-generated assessments.

## 1. Introduction

Sudden cardiac death (SCD) and pump failure death (PFD) are two major causes of mortality in heart failure [1, 2, 3]. Despite advances in pharmacologic therapy and medical devices, accurately identifying individuals at elevated risk for these outcomes remains a persistent clinical challenge [2, 3, 4, 5].

Ambulatory electrocardiograms (ECG), including extended Holter monitoring, supports continuous assessment of cardiac rhythm, capturing transient arrhythmias, ectopic burden, and intermittent conduction abnormalities [6, 7, 8]. These transient abnormalities carry important prognostic information for assessing the risk of SCD and PFD [2, 6, 7, 8]. However, translating ambulatory ECG recordings into reliable and clinically actionable predictors of long-term cardiac mortality remains complex and resource-intensive in routine practice [4, 9, 10].

Machine learning (ML) and artificial intelligence (AI) models offer early promise in addressing these challenges. AI/ML models have demonstrated strong capability in characterizing waveform morphology (e.g., P waves, QRS duration) and identify arrhythmic events (e.g., atrial fibrillation, atrial flutter) [11, 12, 13]. In parallel, large language models (LLMs) have demonstrated capability of reasoning over both structured and unstructured datasets [14, 15, 16]. Domain-adapted medical LLMs can synthesize clinical features (e.g., demographics, medical history, laboratory values) and generate biomedical reasoning [17, 18].

Despite these advancements, important gaps remain.Many AI/ML models for ambulatory ECG analysis are unimodal, focusing primarily on identifying patterns within the ECG recording without incorporating broader patient context [19, 20]. Prior studies suggest that clinical context can influence ECG interpretation by both physicians and AI/ML models [21, 22, 23, 24, 25]. Integrating ECG recordings with patient-level clinical information may therefore provide complementary information for outcome prediction. However, multimodal integration does not necessarily improve performance: its value may depend on the outcome, the representation of each modality, the degree of information overlap, and the fusion strategy. Rigorous comparisons against unimodal and conventional structured-data models are consequently needed to establish when multimodal approaches provide meaningful gains and when they do not.

To address this need, we developed and systematically evaluated a multimodal framework combining patient-level representations derived from ambulatory ECG recordings with clinical information represented as structured variables, deterministic text, or LLM-generated assessments. We used four-year prediction of sudden cardiac death (SCD) and pump failure death (PFD) as our clinical use case. We benchmarked unimodal and multimodal approaches under a common nested cross-validation framework and examined how performance varied across outcomes and clinical representations.

The main contributions of this study are as follows:

1. A rigorous benchmark of ambulatory ECG, tabular clinical variables, deterministic clinical text, and LLM-generated clinical assessments for four-year SCD and PFD prediction, including both unimodal models and ECG–clinical fusion models.
2. A nested cross-validation framework in which multimodal fusion strategies, including concatenation, projected concatenation, scalar gating, vector gating, and weighted sum, were compared and selected using training data only, enabling evaluation of multimodal integration while mitigating model-selection bias.
3. A set of representation and robustness analyses designed to distinguish complementary multimodal information from restatement or duplication of structured variables, including comparisons with conventional tabular models, deterministic text, reasoning and risk-label ablations, ECG-overlap analyses, patient–text shuffling, and attribution stability and perturbation analyses.

## 2. Related Works

### 2.1. Ambulatory ECG Analysis

Early computational studies subsequently emphasized time- and frequency-domain feature extraction to associate ambulatory ECG patterns with cardiac phenotypes. Badilini et al. [26] applied spectral analysis to evaluate circadian variation and beta-blockade effects, while Bakhshi et al. [27] used second-order autoregressive modeling and linear classification to distinguish arrhythmias from sinus rhythm.

More recent work has adopted deep learning to automate representation learning and rhythm classification. Huang et al. [28] transformed ECG segments into spectrograms and applied two-dimensional convolutional neural networks for rhythm classification, whereas Biton et al. [29] proposed ArNet2 to model temporal dependencies across RR intervals. Ben-Moshe et al. [30] introduced RawECGNet to address distributional shifts and detect atrial fibrillation directly from 30-second recordings. Han et al. [31] combined Gramian Angular Field representations with multiple instance learning for arrhythmia detection. Sameh et al. [32] explored how varying the temporal context and front-end representation of ambulatory ECGs affect atrial fibrillation and atrial flutter detection.

Across these approaches, ambulatory ECG recordings are typically analyzed as collections of shorter segments, with predictions generated at the segment level and, in some cases, aggregated through mechanisms such as multiple instance learning to obtain recording-level outputs. However, these strategies are often limited to localized rhythm detection.

### 2.2. LLMs and Multimodal Integration

LLMs and domain-adapted language models have been studied for their utility in understanding clinical text and decision-support applications. Lee et al. introduced BioBERT [33], and Huang et al. developed ClinicalBERT [34], both leveraging domain-specific pretraining to adapt transformer architectures to biomedical and clinical corpora for improved encoding of clinical narratives and structured electronic health record data. More recently, Xie et al. proposed Me-LLaMA [17], and Li et al. introduced LLaVA-Med [35], exploring instruction tuning and multimodal alignment strategies to enable higher-level clinical reasoning, including summarization, question answering, and outcome prediction from heterogeneous patient information. Yang et al. [36] constructed ECG-LM, which processes both short-duration ECGs and clinical notes for classifying arrhythmic events and propose care strategies.

These approaches apply language models to textual data and, in some cases, short-duration ECG signals, primarily focusing on representation learning and arrhythmia classification tasks. To our knowledge, there has been limited work in integrating patient clinical context with 24-hour ambulatory ECG signals.

## 3. Methods

Fig. 1 illustrates our multimodal workflow for analyzing ambulatory ECG recordings together with clinical features. ECG recordings are preprocessed and segmented into short windows. For each window, heart rate variability (HRV) features are extracted and encoded via a Multiple Instance Learning–Temporal Convolutional Network (MIL–TCN), generating an ECG representation, 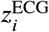. In parallel, clinical variables are represented as LLM-generated text, deterministic clinical text, or tabular features. The LLM-generated and deterministic text are encoded via a biomedical text encoder to produce patient-level text representations, 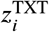, whereas the preprocessed tabular variables form the tabular representation, 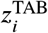.

**Figure 1:**
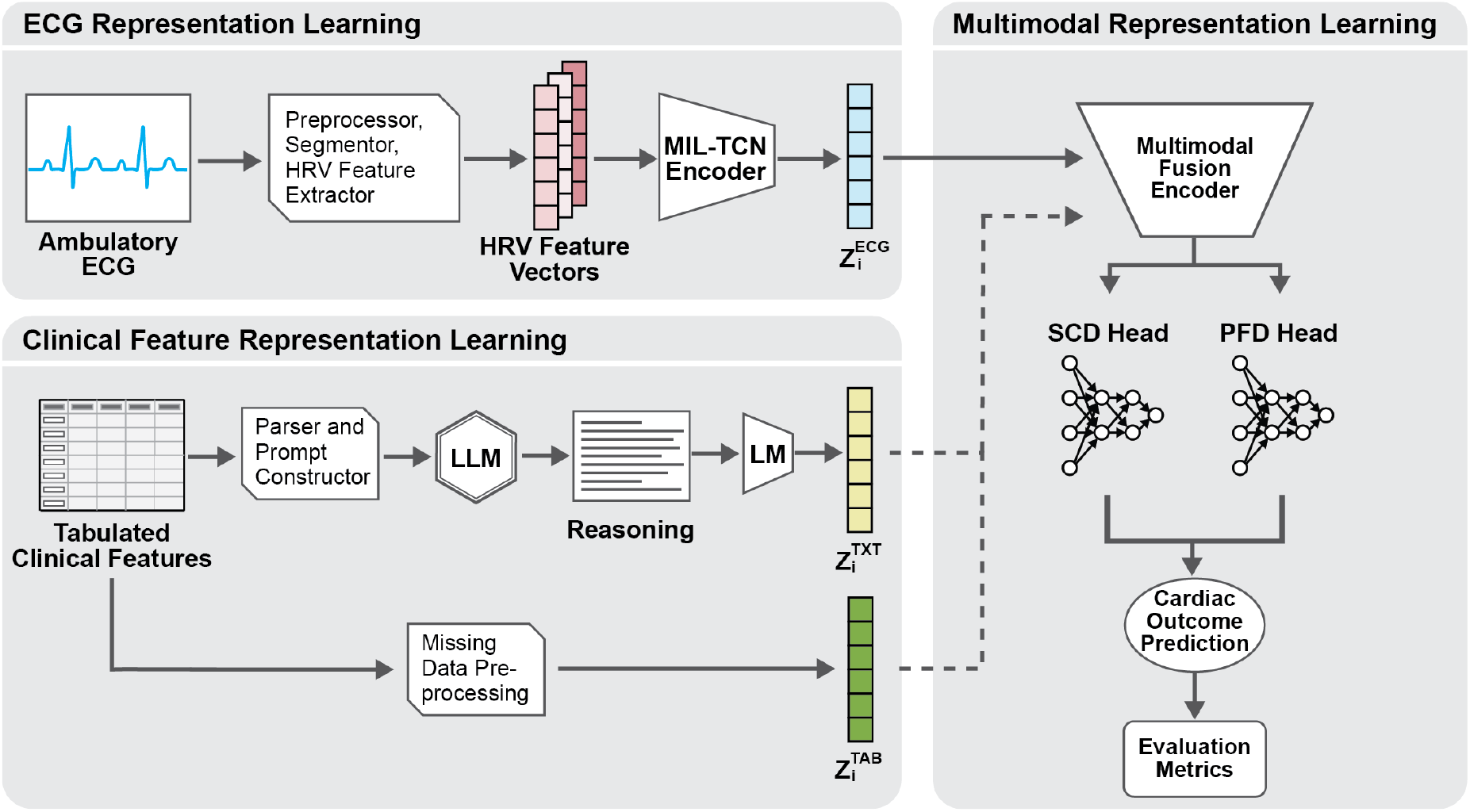
The Multimodal Workflow for Ambulatory ECG and Clinical Feature Analysis. Dashed lines indicate using either the text (TXT) or tabular (TAB) representation for multimodal fusion.

The ECG, text, and tabular representations were each evaluated as unimodal models, while the text and tabular representations were also paired separately with the ECG representation in multimodal models for SCD and PFD prediction.

### 3.1. ECG Representation Learning

#### 3.1.1. ECG Preprocessing

All ambulatory ECG recordings underwent a standardized preprocessing pipeline for denoising. Signals were bandpass filtered between 0.5–40 Hz using a fourth-order Butterworth filter to attenuate baseline wander and high-frequency noise [37]. Baseline drift was further corrected via median filtering over a 0.8-second window, and the estimated baseline component was subtracted from the filtered signal [38]. R-peaks were then detected once per full-length recording using NeuroKit2 to ensure consistent beat localization across the entire ambulatory trace [39]. Full-record NeuroKit2 processing was subsequently performed to derive physiologic descriptors, including instantaneous heart rate and signal-quality indices.

Each processed recording was divided chronologically into non-overlapping 30-second segments. Only complete segments were retained. Within each segment, time-domain heart rate variability (HRV) measures, including SDNN, RMSSD, and pNN50, were computed. The resulting segment-level features were stored as feature vectors for downstream modeling. A detailed table of the computed features can be found in Table A.1 in Appendix A.

#### 3.1.2. Multiple Instance Learning with Temporal Convolutional Encoding

Under the multiple instance learning (MIL) paradigm [40, 41], each patient recording was represented as a bag of chronologically ordered segment-level instances. Each patient *i* was assigned binary outcome indicators for sudden cardiac death (SCD) and pump failure death (PFD), denoted by 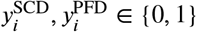

Each patient’s ambulatory ECG was represented by *N*_*i*_ non-overlapping 30-second segments, yielding a bag

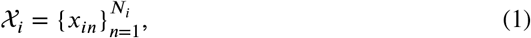

where *x*_*in*_ ∈ ℝ^*F*^ denotes the ECG-derived feature vector for segment *n* and *F* is the number of input features after preprocessing. The number of segments, *N*_*i*_, varied across patients according to recording duration and the number of complete segments.

Each segment was first projected into a learned embedding space using a shared feature encoder *f* _*θ*_: ℝ^*F*^ →ℝ^*d*^

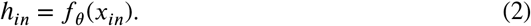

To model temporal dependencies across segments, the ordered segment embeddings were assembled as

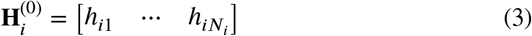

and processed by a residual temporal convolutional network (TCN). The *l* ^th^block applied a one-dimensional causal convolution with dilation 2^*l*^, followed by a rectified linear unit and dropout:

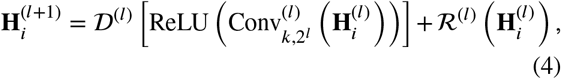

where *k* is the convolutional kernel size, *D*^(*l*)^ denotes dropout, and ℛ ^(*l*)^ is either the identity mapping or a learned projection when the input and output dimensions differ. Padding followed by right truncation preserved the original sequence length. The output corresponding to segment *n* after the final temporal block is denoted by 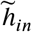.

The variable-length sequence was aggregated into a patient-level vector using attention-based MIL pooling:

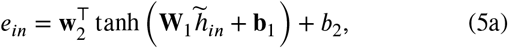

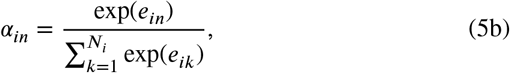

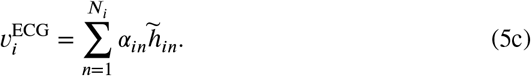

The attention-pooled vector was transformed by a shared patient-level representation layer:

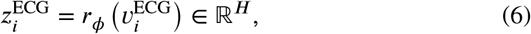

where *H* denotes the selected ECG representation dimension. The resulting ECG representation, 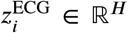, was passed to separate task-specific branches for SCD and PFD prediction.

### 3.2. Clinical Feature Representation via Language Models

#### 3.2.1 Prompt Construction

We utilized two instruction-tuned large language models, LLaMA-3.1-8B-Instruct and LLaMA-3.2-3B-Instruct [42, 43]. These LLMs were used to generate clinical assessments from patient clinical features. Clinical variables were parsed and inserted into a standardized template describing demographics, comorbidities, medications, laboratory measurements, and, for the designated ECG-inclusive condition, summary ambulatory ECG impressions. Variables with missing baseline values were omitted from the patient-specific text rather than imputed or inferred by the LLM. Each prompt contained the complete task instructions and patient-specific clinical features; no additional system message containing clinical instructions was added during inference. The full-risk prompts instructed the model to independently assess the four-year risk of SCD and PFD. The model was required to produce categorical risk estimates (low, moderate, or high) together with separate explanatory rationales for each outcome.

Three LLM-generated text conditions were evaluated: a full four-year risk assessment without ECG impressions, a neutral clinical summary without ECG impressions, and a full four-year risk assessment with ECG impressions. The ECG-inclusive condition was evaluated only as an ablation and was not used in the primary LLM-text analyses. Risk-label-only and rationale-only representations were subsequently derived from the structured full-risk responses.

For comparison, a deterministic text representation was constructed directly from the populated patient-information template. This representation preserved the same clinical variables in a fixed field order but contained no LLM-generated summary, risk estimate, or rationale, serving as a control for representing the structured variables in textual form. No outcome labels or post-index follow-up information were included in the prompts. All prompt templates can be found in Appendix B.

#### 3.2.2. Text Embedding Generation

Let the text input for patient *i*, consisting of either LLM-generated clinical text or deterministic text, be denoted by *t*_*i*_. Each text input was encoded using a frozen biomedical text encoder, either BioBERT [33] or ClinicalBERT [34], to produce a fixed-length embedding *e*_*i*_ ∈ ℝ^*p*^ obtained from the final hidden representation of the `[CLS]` token. These embeddings served as patient-level representations of the generated or deterministic clinical text. Each embedding was then passed through a feature transformation 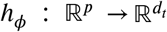, implemented as either a linear layer or a multilayer perceptron, to produce the patient-level representation

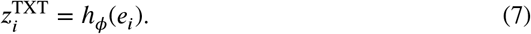

The representation 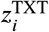 was then passed to task-specific branches for SCD and PFD prediction.

### 3.3. Tabular Clinical-Feature Models

To assess whether converting structured clinical variables into text preserved or improved their predictive value, the same variables were modeled directly in tabular form. The primary tabular condition included the baseline demographics, clinical characteristics, medical history, laboratory measurements, cardiac function, and medications used to construct the LLM prompts without ECG information. A second condition additionally included the five Holter-derived ECG impressions used in the ECG-inclusive prompt: ventricular extrasystole, ventricular tachycardia, non-sustained ventricular tachycardia, paroxysmal supraventricular tachyarrhythmia, and bradycardia. This ECG-inclusive tabular condition was evaluated only as an ablation and was not used in the primary tabular analyses.

For missing data imputation, continuous variables were replaced by their median values and supplemented with missingness indicators, whereas missing binary and categorical values were represented as a separate category. Continuous variables were subsequently standardized to have a mean of zero and a standard deviation of one, and binary and categorical variables were one-hot encoded. All preprocessing parameters were estimated using only the data available for model fitting and applied unchanged during evaluation. Additionally, two conventional tabular model families were evaluated: L2-penalized logistic regression and histogram-based gradient boosting. Separate models were constructed for SCD and PFD prediction.

### 3.4. Multimodal Fusion Architecture

#### 3.4.1. Preparation of ECG, Text, and Tabular Representations

Let *d*_*e*_, *d*_*t*_, and *d*_*c*._ denote the dimensions of the ECG, text, and tabular representations, respectively. For the ECG modality, the trained MIL–TCN encoder was frozen, and the patient-level representation 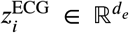derived from the attention-pooled output was extracted for each patient. For the text modality, embeddings 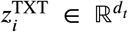 were ob-tained from the frozen biomedical text encoder. These included embeddings of the LLM-generated risk assessments and deterministic clinical text. For the tabular modality, 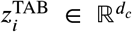was the preprocessed clinical-feature vector comprising standardized continuous variables, continuous-variable missingness indicators, and one-hot-encoded categorical variables, as described above. The tabular representation was used directly rather than extracted from a trained tabular classifier. Moreover, the ECG representation was paired separately with the LLM-generated text, deterministic text, and tabular clinical representations. These modality-specific representations served as inputs to the multimodal fusion models described below.

#### 3.4.2. Multimodal Fusion Strategies

Given the ECG representation 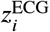and text representation 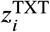 for patient *i*, we evaluated five multimodal fusion strategies to produce a fused representation 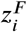. The same strategies were used for ECG–tabular fusion by replacing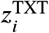 with the corresponding preprocessed tabular represen-tation 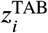.

**Direct concatenation (DC)** concatenates the modality-specific representations without projection:

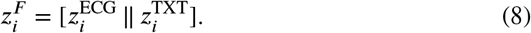

**Projected concatenation (PC)** first maps each modality into a shared latent space:

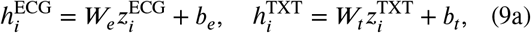

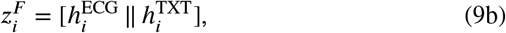

where *W*_*e*_ and, *W*_*t*_ are learned projection matrices.

**Scalar gating (SG)** uses a patient-specific scalar gate conditioned on both projected representations:

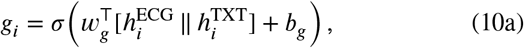

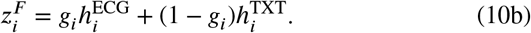

**Vector gating (VG)** computes a patient-specific gating vector conditioned on both modalities:

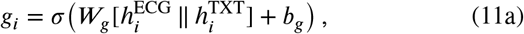

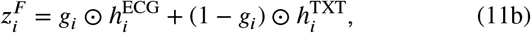

where ⊙denotes element-wise multiplication.

**Weighted sum (WS)** combines the projected representations using a single global learned mixture weight:

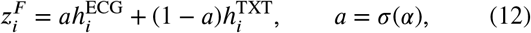

where α ∈ ℝ is a learned scalar parameter shared across patients.

In all cases, 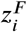was passed to separate task-specific prediction branches for SCD and PFD.

For interpretation of the ECG + LLM text multimodal models, ECG importance was assessed using MIL attention weights, while text importance was assessed using gradient-based token saliency. For each token, saliency was computed as the Euclidean norm of the element-wise product between its embedding and the gradient of the predicted SCD or PFD probability with respect to that embedding. Token scores were mapped back to the LLM-generated clinical text for visualization and therefore reflect the text pathway within the fitted multimodal model rather than the internal attention weights of the LLaMA generator.

### 3.5. Model Training and Configurations

#### 3.5.1. Loss Functions

The ECG, text, and multimodal fusion networks were trained using a multitask objective to jointly predict SCD and PFD. For each task, we used weighted binary cross-entropy loss, with the positive-class weight set to the ratio of negative to positive cases in the training data:

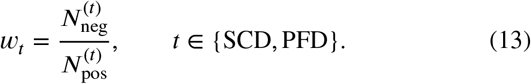

Patients with PFD were omitted from the SCD loss, and patients with SCD were omitted from the PFD loss, so that competing cardiac-death events were not treated as negative outcomes. Patients without cardiac death contributed as negative cases to both tasks. The overall objective was the sum of the two task-specific losses:

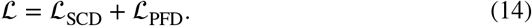

The conventional tabular models were fitted separately for SCD and PFD using the same task-specific class-weighting approach and exclusion of the competing cardiac-death outcome.

#### 3.5.2. LLM and Biomedical Text Encoder Configurations

Clinical text outputs were generated using Hugging Face Transformers v4.44.2 with two instruction-tuned models: LLaMA-3.1-8B-Instruct and LLaMA-3.2-3B-Instruct. The LLaMA models were loaded in 16-bit floating-point precision without quantization. Generation used deterministic greedy decoding with `do_sample=False,` a maximum of 256 new tokens, a repetition penalty of 1.1, a batch size of four, and a generation seed of 42. Temperature, top-*p*, and top-*k* were not applicable because sampling was disabled. Generation was performed using four NVIDIA GeForce RTX 2080 Ti GPUs with balanced device placement (see Section 3.5.3 for hardware specifications). All LLM inference was performed locally using downloaded model weights; patient-level data were not transmitted to an external commercial LLM service.

Generated and deterministic text inputs were encoded using frozen BioBERT or ClinicalBERT biomedical text encoders. The encoders were kept frozen to limit the number of trainable parameters relative to the cohort and event counts and to reduce the risk of overfitting during nested model selection. This design also allowed comparisons to focus on differences arising from the clinical-text representations rather than outcome-specific adaptation of the biomedical text encoder. Text was processed with a maximum sequence length of 512 tokens. For inputs exceeding this length, overlapping 512-token chunks with a stride of 64 tokens were encoded separately, and the final hidden representations of the `[CLS]` tokens were averaged across chunks to obtain a single 768-dimensional text embedding. The two LLaMA generators and two biomedical text encoders defined four candidate LLaMA–encoder combinations. Their corresponding model hashes are reported in Table B.1 in Appendix B.

#### 3.5.3. Training Protocol, Cross-Validation, and Model Selection

All experiments were conducted on a Linux server running Ubuntu with NVIDIA GeForce RTX 2080 Ti GPUs. Neural models were implemented in PyTorch v2.5.1 with CUDA 12.4. Models were developed and evaluated using patient-level nested cross-validation [44, 45], with five mutually exclusive outer folds and four inner folds and a base random seed of 42. The folds were stratified by the mutually exclusive four-year outcome categories of no cardiac death, SCD, and PFD. All ECG segments, text representations, and tabular clinical representations from a given patient were assigned to the same fold.

Within each outer training fold, four-fold inner cross-validation was used for all model-development steps, including preprocessing, feature scaling, class weighting, hyperparameter and model-configuration selection, early stopping, fusion-strategy selection, probability calibration, and classification-threshold selection. After model selection, the selected configuration was fitted using the complete outer training fold and evaluated once on the corresponding outer-test fold. The outer-test fold remained unseen during model development, and its outcomes were not used for preprocessing, tuning, calibration, threshold selection, or model selection. Details of variable provenance and hyperparameter search spaces are provided in Appendix C and Appendix D, respectively.

Early-stopping settings were fixed across candidate configurations rather than treated as tunable hyperparameters. ECG models were trained for a maximum of 30 epochs, with a minimum of five epochs and a patience of seven epochs. Text classifiers and multimodal fusion models were trained for a maximum of 100 epochs, with a minimum of 10 epochs and a patience of 15 epochs. The checkpoint with the highest mean validation AUROC across the SCD and PFD tasks was retained.

For each candidate configuration, predictions from the inner validation folds were pooled before calculating the SCD and PFD AUROCs. Model selection was based on the mean of these two AUROCs, with the endpoints weighted equally. Configurations within an absolute AUROC differ-ence of 0.005 from the highest-performing configuration were compared by model complexity and then by mean PR-AUC. For neural models, complexity was defined by the number of trainable parameters. For the tabular models, a prespecified ordering favored penalized logistic regression over gradient boosting and favored simpler, more strongly regularized configurations within each model family.

## 4. Experiments

### 4.1. Dataset and Patient Cohort Selection

Experiments were conducted using the publicly available MUSIC (MUerte S’ubita en Insuficiencia Cardiaca) dataset, a prospective multicenter cohort of patients with chronic heart failure [46, 47, 48]. The dataset contains deidentified baseline clin information, including demographic characteristics, medical history, laboratory measurements, medications, and cardiac-function measures. Only the deidentified variables available in the public dataset were used in this study; no direct patient identifiers were included in the LLM prompts. The study enrolled 992 patients from eight Spanish university hospitals between April 2003 and December 2004, with a median follow-up of 44 months. Ambulatory ECG data were acquired using SpiderView recorders (ELA Medical, Sorin Group, Paris, France) with two or three orthogonal leads (X, Y, and Z) sampled at 200 Hz with 10 μV amplitude resolution. Only Lead Y was used for signal analysis. This lead corresponds to the superior–inferior electrical axis and has an orientation similar to Lead II of the standard 12-lead ECG, making it suitable for beat detection and heart rate variability analysis using NeuroKit2 [39, 49].

The baseline 24-hour Holter recording was used as the index time, and outcomes were defined over a fixed four-year prediction horizon. Cohort selection is summarized in Fig. 2, with corresponding baseline characteristics in Table 1 and ECG impressions in Table 2. Starting from the 992 enrolled patients, we excluded 56 without an available Holter recording; 57 whose recorded outcome fell outside the prespecified categories of no cardiac death, SCD, and PFD; 121 with a prior implantable cardiac device; 12 who exited the study because of cardiac transplantation; and 16 whose four-year outcome could not be ascertained. The two dataset outcome codes representing PFD were combined into a single category, and SCD and PFD were mutually exclusive within the prediction horizon. The final cohort included 730 patients with three mutually exclusive four-year outcome labels: 577 with no cardiac death, 71 with SCD, and 82 with PFD. Patient-level nested cross-validation used five mutually exclusive outer folds of 146 patients each, yielding 584 patients for outer-fold model development and 146 patients for held-out evaluation in each iteration. The outer folds were stratified by the three outcome categories, and each outer training set underwent four-fold inner cross-validation for model development and selection. For the separate binary classification tasks, the SCD group was compared with the no-cardiac-death group while patients with PFD were excluded (*N* = 648), and the PFD group was compared with the no-cardiac-death group while patients with SCD were excluded (*N* = 659). Detailed cohort exclusions and endpoint-specific analysis populations are provided in Appendix E.

**Table 1.**
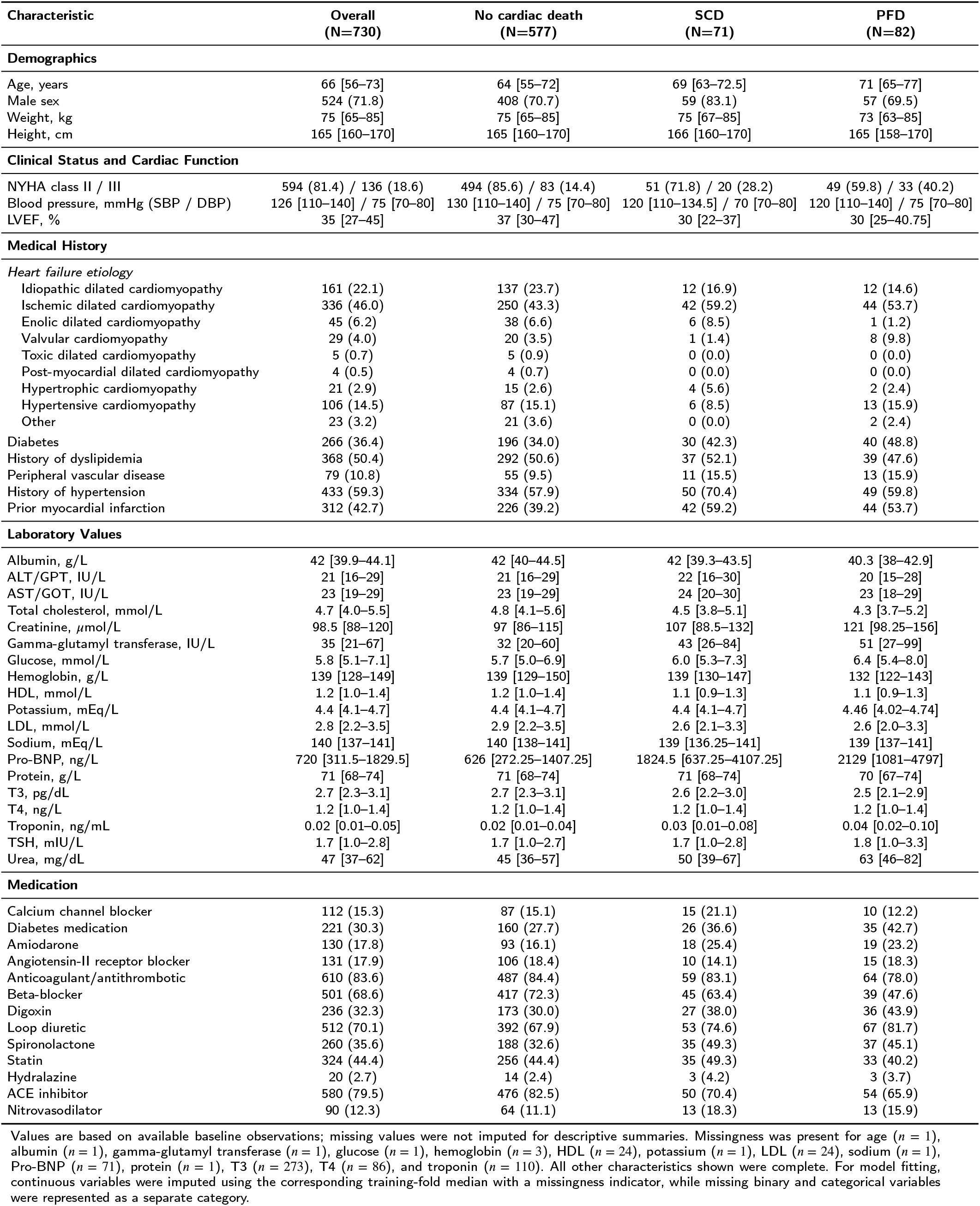
Baseline characteristics of the final four-year MUSIC cohort, stratified by outcome. Continuous variables are reported as median [interquartile range] and categorical variables as n (%).

**Table 2.** Baseline Holter-derived ECG impressions in the final four-year MUSIC cohort, stratified by outcome. Values are reported as n (%).

| ECG impression | Overall<br>(N=730) | No cardiac death<br>(N=577) | SCD<br>(N=71) | PFD<br>(N=82) |
| --- | --- | --- | --- | --- |
| *Ventricular extrasystole* |  |  |  |  |
| No | 19 (2.6) | 18 (3.1) | 0 (0.0) | 1 (1.3) |
| Monomorphic | 213 (29.5) | 187 (32.7) | 11 (15.7) | 15 (18.8) |
| Polymorphic | 490 (67.9) | 367 (64.2) | 59 (84.3) | 64 (80.0) |
| *Ventricular tachycardia* |  |  |  |  |
| No | 460 (63.9) | 387 (68.1) | 35 (50.0) | 38 (46.3) |
| Non-sustained VT | 260 (36.1) | 181 (31.9) | 35 (50.0) | 44 (53.7) |
| *Non-sustained ventricular tachycardia* |  |  |  |  |
| No | 523 (71.6) | 439 (76.1) | 37 (52.1) | 47 (57.3) |
| Yes | 207 (28.4) | 138 (23.9) | 34 (47.9) | 35 (42.7) |
| *Paroxysmal supraventricular tachyarrhythmia* |  |  |  |  |
| No | 406 (56.2) | 332 (58.3) | 35 (49.3) | 39 (47.6) |
| TPSV | 168 (23.3) | 135 (23.7) | 16 (22.5) | 17 (20.7) |
| Paroxysmal AF | 4 (0.6) | 2 (0.4) | 1 (1.4) | 1 (1.2) |
| Paroxysmal atrial flutter | 1 (0.1) | 0 (0.0) | 1 (1.4) | 0 (0.0) |
| Not applicable because of chronic AF | 143 (19.8) | 100 (17.6) | 18 (25.4) | 25 (30.5) |
| *Bradycardia* |  |  |  |  |
| No | 532 (92.4) | 435 (92.8) | 46 (88.5) | 51 (92.7) |
| Sinus node dysfunction | 17 (3.0) | 13 (2.8) | 3 (5.8) | 1 (1.8) |
| First-degree AV block | 8 (1.4) | 5 (1.1) | 1 (1.9) | 2 (3.6) |
| Second-degree AV block, type I | 9 (1.6) | 8 (1.7) | 0 (0.0) | 1 (1.8) |
| Second-degree AV block, type II | 7 (1.2) | 6 (1.3) | 1 (1.9) | 0 (0.0) |
| Third-degree AV block | 1 (0.2) | 0 (0.0) | 1 (1.9) | 0 (0.0) |
| Paroxysmal AV block | 2 (0.3) | 2 (0.4) | 0 (0.0) | 0 (0.0) |

**Figure 2:**
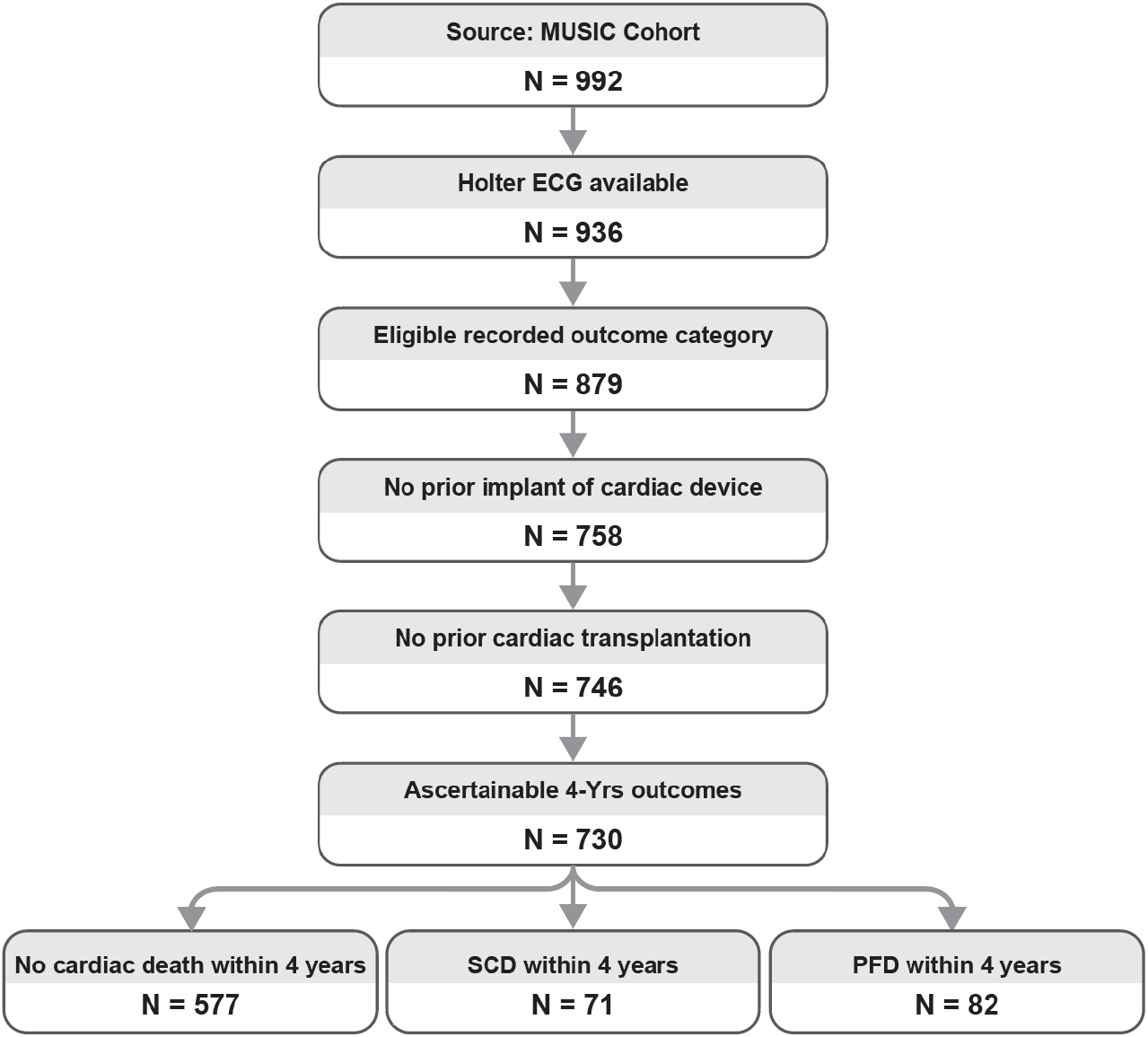
MUSIC cohort selection and four-year outcome groups.

### 4.2. Performance Evaluation

Performance was evaluated using the pooled outer-test predictions. Predictions from all five outer folds were combined separately for the SCD and PFD endpoint cohorts, and each performance metric was calculated once from the corresponding pooled set of held-out predictions. Discrimination was evaluated from the raw prediction scores using the area under the receiver operating characteristic curve (AUROC) and the area under the precision–recall curve (PR-AUC) [50].

Probability calibration was performed separately within each outer fold and outcome using Platt scaling fitted to inner out-of-fold predictions from the outer training cohort. The fitted calibration mapping was applied unchanged to the corresponding outer-test predictions. Calibration was assessed using calibration plots, calibration intercept, calibration slope, and the Brier score. Calibration intercept and slope were estimated by regressing the observed binary outcome on the log-odds of the calibrated predicted probability:

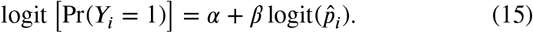

where 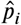 is the calibrated predicted probability for patient *i* for the positive outcome (i.e., SCD or PFD). The Brier score was calculated as

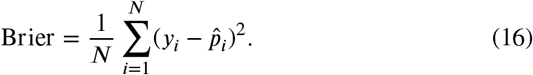

For threshold-specific evaluation, a classification threshold was selected separately for each outer fold and endpoint by maximizing Youden’s *J* statistic using only the calibrated inner out-of-fold predictions. Ties were resolved by selecting the threshold with higher sensitivity and then the lower threshold. The selected threshold was applied unchanged to the corresponding outer-test predictions. Let *TP, TN, FP*, and *FN* denote the numbers of true positives, true negatives, false positives, and false negatives, respectively. The following threshold-specific metrics were calculated:

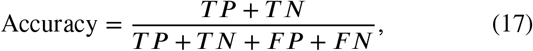

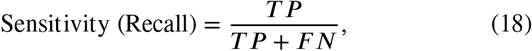

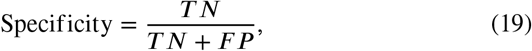

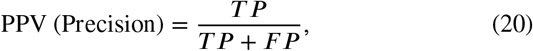

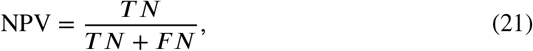

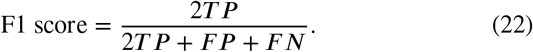

Uncertainty was quantified using 5,000 patient-level bootstrap replicates, from which 95% confidence intervals were calculated for discrimination, calibration, and threshold-specific metrics. Comparisons between models used paired bootstrap resampling, with the same resampled patients used for both models in each replicate. Holm’s procedure was used to adjust for multiple comparisons within each prespecified comparison family. A p-value less than 0.05 was considered statistically significant.

Exploratory decision curve analysis was performed using the fold-specific calibrated outer-test probabilities [51]. At a risk threshold *p*_*i*_, net benefit was calculated as

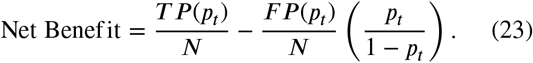

Model net benefit was compared with treat-all and treat-none strategies over thresholds from 2% to 25%, with pointwise patient-level bootstrap uncertainty. This exploratory range encompassed the observed event prevalences and a broad range of potential risk tolerances; however, no intervention-specific threshold range has been clinically validated for predicted four-year SCD or PFD risk [52, 53]. Accordingly, the decision curves were used to compare model behavior and were not interpreted as evidence of established clinical utility.

### 4.3. Experimental Results

The performance metrics for SCD and PFD prediction for both unimodal and multimodal models are summarized in Table 3 and Table 4, respectively. Corresponding confusion matrices, AUROC curves, PR-AUC plots, and exploratory decision curve plots are shown in Fig. 3, Fig. 4, Fig. 5, and Fig. 6, respectively. Corresponding calibration plots and tables are in Figure F.1 and Table F.1 in Appendix F. Detailed model comparisons are shown in Table F.2 and Table F.3, also in Appendix F. In addition to quantitative performance evaluation, Fig. 7 and Fig. 8 visualize multimodal model attribution across ECG segments and clinical text for representative SCD and PFD cases. Attribution robustness was evaluated using repeated model refitting, meaning-preserving input perturbations, and token-masking experiments. Representation and overlap ablations compared LLM-generated text, deterministic text, direct tabular clinical variables, and the inclusion of Holter-derived ECG impressions, as summarized in Table 5.

**Table 3.** Comparison of unimodal and multimodal models for four-year sudden cardiac death prediction. Results are pooled outer-test estimates with 95% confidence intervals. Multimodal results use the fusion architecture selected independently within each outer-training fold using inner cross-validation. Bold indicates the best point estimate.

| Model | AUROC | PR-AUC | Brier score | Accuracy | PPV/Precision | Sensitivity/Recall | Specificity | NPV | F1 |
| --- | --- | --- | --- | --- | --- | --- | --- | --- | --- |
| **Unimodal** |  |  |  |  |  |  |  |  |  |
| ECG | 0.628<br>(0.548–0.710) | 0.212<br>(0.149–0.314) | 0.097<br>(0.079–0.116) | **0.775**<br>(0.742–0.807) | **0.218**<br>(0.149–0.292) | 0.408<br>(0.297–0.529) | **0.820**<br>(0.787–0.850) | 0.918<br>(0.894–0.941) | 0.284<br>(0.201–0.366) |
| Full LLM text | 0.531<br>(0.457–0.603) | 0.132<br>(0.093–0.187) | 0.100<br>(0.082–0.119) | 0.628<br>(0.591–0.665) | 0.121<br>(0.079–0.165) | 0.380<br>(0.266–0.493) | 0.659<br>(0.619–0.696) | 0.896<br>(0.867–0.924) | 0.183<br>(0.123–0.243) |
| Deterministic text | 0.681<br>(0.615–0.745) | 0.204<br>(0.150–0.298) | 0.094<br>(0.078–0.112) | 0.523<br>(0.486–0.563) | 0.152<br>(0.116–0.191) | **0.732**<br>(0.627–0.831) | 0.497<br>(0.457–0.540) | 0.938<br>(0.909–0.964) | 0.252<br>(0.197–0.307) |
| Tabular clinical features | 0.696<br>(0.625–0.764) | 0.244<br>(0.175–0.352) | 0.093<br>(0.076–0.111) | 0.650<br>(0.613–0.685) | 0.188<br>(0.139–0.236) | 0.662<br>(0.548–0.769) | 0.648<br>(0.609–0.685) | **0.940**<br>(0.914–0.961) | 0.293<br>(0.226–0.356) |
| **Multimodal** |  |  |  |  |  |  |  |  |  |
| ECG + full LLM text | 0.591<br>(0.517–0.665) | 0.158<br>(0.117–0.234) | 0.098<br>(0.080–0.119) | 0.660<br>(0.623–0.696) | 0.153<br>(0.108–0.204) | 0.465<br>(0.349–0.580) | 0.685<br>(0.646–0.722) | 0.912<br>(0.884–0.937) | 0.231<br>(0.167–0.295) |
| ECG + deterministic text | **0.711**<br>(0.643–0.773) | 0.242<br>(0.176–0.348) | **0.091**<br>(0.075–0.110) | 0.733<br>(0.699–0.765) | 0.213<br>(0.154–0.279) | 0.535<br>(0.412–0.650) | 0.757<br>(0.721–0.791) | 0.930<br>(0.905–0.953) | **0.305**<br>(0.227–0.382) |
| ECG + tabular clinical features | 0.689<br>(0.611–0.760) | **0.248**<br>(0.180–0.357) | 0.092<br>(0.074–0.111) | 0.713<br>(0.677–0.747) | 0.202<br>(0.148–0.265) | 0.549<br>(0.435–0.667) | 0.733<br>(0.696–0.768) | 0.930<br>(0.904–0.952) | 0.295<br>(0.225–0.370) |

**Table 4.** Comparison of unimodal and multimodal models for four-year pump failure death prediction. Results are pooled outer-test estimates with 95% confidence intervals. Multimodal results use the fusion architecture selected independently within each outer-training fold using inner cross-validation. Bold indicates the best point estimate.

| Model | AUROC | PR-AUC | Brier score | Accuracy | PPV/Precision | Sensitivity/Recall | Specificity | NPV | F1 |
| --- | --- | --- | --- | --- | --- | --- | --- | --- | --- |
| **Unimodal** |  |  |  |  |  |  |  |  |  |
| ECG | 0.628<br>(0.566–0.687) | 0.182<br>(0.137–0.255) | 0.107<br>(0.089–0.125) | 0.531<br>(0.492–0.568) | 0.169<br>(0.130–0.210) | 0.707<br>(0.607–0.805) | 0.506<br>(0.465–0.546) | 0.924<br>(0.894–0.951) | 0.273<br>(0.216–0.328) |
| Full LLM text | 0.467<br>(0.399–0.536) | 0.131<br>(0.096–0.192) | 0.111<br>(0.093–0.131) | 0.524<br>(0.486–0.563) | 0.118<br>(0.083–0.156) | 0.439<br>(0.333–0.544) | 0.536<br>(0.496–0.579) | 0.870<br>(0.835–0.904) | 0.187<br>(0.133–0.239) |
| Deterministic text | 0.680<br>(0.611–0.746) | 0.300<br>(0.214–0.400) | 0.102<br>(0.085–0.119) | 0.675<br>(0.639–0.712) | 0.211<br>(0.158–0.265) | 0.585<br>(0.480–0.690) | 0.688<br>(0.649–0.726) | 0.921<br>(0.895–0.946) | 0.310<br>(0.241–0.375) |
| Tabular clinical features | **0.787**<br>(0.732–0.839) | **0.360**<br>(0.275–0.470) | **0.096**<br>(0.079–0.113) | 0.737<br>(0.704–0.771) | 0.288<br>(0.230–0.352) | **0.756**<br>(0.658–0.849) | 0.735<br>(0.699–0.770) | **0.955**<br>(0.935–0.973) | **0.418**<br>(0.345–0.489) |
| **Multimodal** |  |  |  |  |  |  |  |  |  |
| ECG + full LLM text | 0.603<br>(0.538–0.666) | 0.170<br>(0.130–0.240) | 0.109<br>(0.091–0.128) | 0.646<br>(0.610–0.681) | 0.184<br>(0.136–0.236) | 0.537<br>(0.431–0.643) | 0.662<br>(0.623–0.699) | 0.910<br>(0.882–0.936) | 0.274<br>(0.209–0.338) |
| ECG + deterministic text | 0.675<br>(0.612–0.735) | 0.240<br>(0.172–0.327) | 0.104<br>(0.086–0.122) | 0.677<br>(0.640–0.713) | 0.209<br>(0.156–0.264) | 0.573<br>(0.463–0.681) | 0.692<br>(0.653–0.728) | 0.919<br>(0.892–0.944) | 0.306<br>(0.236–0.373) |
| ECG + tabular clinical features | 0.756<br>(0.697–0.811) | 0.353<br>(0.261–0.461) | 0.097<br>(0.080–0.114) | **0.763**<br>(0.730–0.794) | **0.292**<br>(0.227–0.361) | 0.634<br>(0.529–0.741) | **0.782**<br>(0.747–0.814) | 0.938<br>(0.915–0.958) | 0.400<br>(0.323–0.476) |

**Table 5.** Representation and overlap ablations for four-year sudden cardiac death and pump failure death prediction. Results are pooled outer-test estimates with 95% confidence intervals. Bold indicates the best point estimate.

| Representation | SCD |  |  | PFD |  |  |
| --- | --- | --- | --- | --- | --- | --- |
|  | AUROC | PR-AUC | Brier score | AUROC | PR-AUC | Brier score |
| **LLM-generated and deterministic text representations** |  |  |  |  |  |  |
| Full LLM assessment, no ECG impressions | 0.531<br>(0.463–0.601) | 0.130<br>(0.093–0.185) | 0.100<br>(0.082–0.119) | 0.459<br>(0.388–0.528) | 0.134<br>(0.096–0.198) | 0.111<br>(0.092–0.131) |
| Risk label only, no ECG impressions | 0.503<br>(0.433–0.574) | 0.111<br>(0.086–0.151) | 0.098<br>(0.079–0.116) | 0.494<br>(0.430–0.562) | 0.122<br>(0.096–0.156) | 0.109<br>(0.091–0.129) |
| Rationale only, no ECG impressions | 0.517<br>(0.442–0.589) | 0.129<br>(0.092–0.186) | 0.099<br>(0.081–0.118) | 0.490<br>(0.422–0.558) | 0.140<br>(0.104–0.206) | 0.110<br>(0.091–0.129) |
| Neutral clinical summary, no ECG impressions | 0.574<br>(0.503–0.645) | 0.141<br>(0.103–0.202) | 0.099<br>(0.081–0.118) | 0.620<br>(0.553–0.686) | 0.188<br>(0.141–0.260) | 0.107<br>(0.089–0.126) |
| Deterministic non-LLM template | 0.681<br>(0.615–0.745) | 0.204<br>(0.150–0.298) | 0.094<br>(0.078–0.112) | 0.680<br>(0.611–0.746) | 0.300<br>(0.214–0.400) | 0.102<br>(0.085–0.119) |
| Full LLM assessment, with ECG impressions | 0.516<br>(0.446–0.588) | 0.117<br>(0.088–0.170) | 0.098<br>(0.080–0.117) | 0.655<br>(0.589–0.722) | 0.245<br>(0.176–0.335) | 0.105<br>(0.088–0.124) |
| **Tabular clinical-variable controls** |  |  |  |  |  |  |
| Penalized logistic regression, clinical variables only | 0.696<br>(0.625–0.764) | 0.244<br>(0.175–0.352) | 0.093<br>(0.076–0.111) | 0.787<br>(0.732–0.839) | 0.360<br>(0.275–0.470) | **0.096**<br>(0.079–0.113) |
| Penalized logistic regression, clinical variables with ECG impressions | **0.710**<br>(0.643–0.774) | **0.252**<br>(0.182–0.359) | **0.092**<br>(0.076–0.110) | **0.792**<br>(0.736–0.842) | **0.364**<br>(0.273–0.476) | **0.096**<br>(0.080–0.113) |

**Figure 3:**
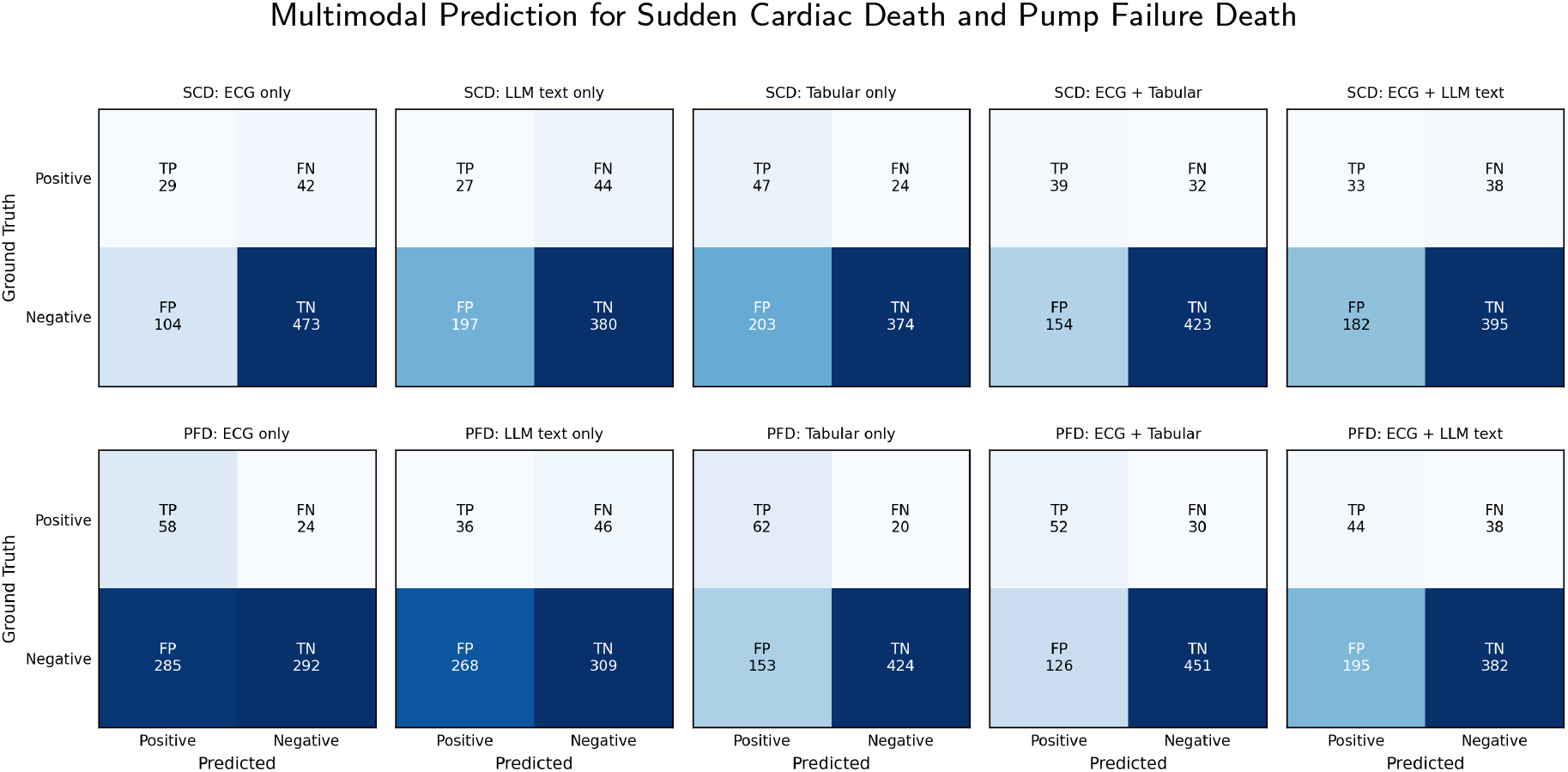
Comparison of confusion matrices for unimodal (ECG only, LLM text only, Tabular only) and multimodal (ECG + Tabular, ECG + LLM Text) models for sudden cardiac death (a) and pump failure death (b).

**Figure 4:**
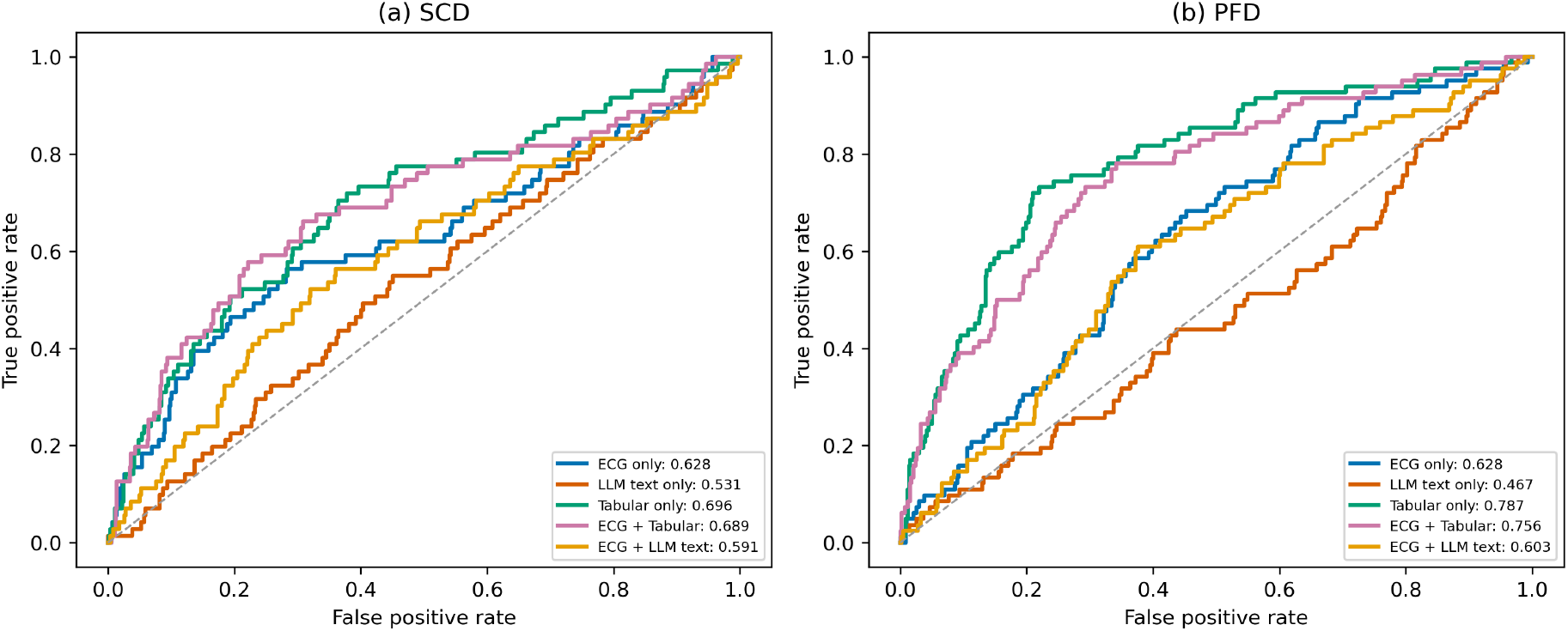
Comparison of AUROC curves for unimodal (ECG only, LLM text only, Tabular only) and multimodal (ECG + Tabular, ECG + LLM Text) models for sudden cardiac death (a) and pump failure death (b).

**Figure 5:**
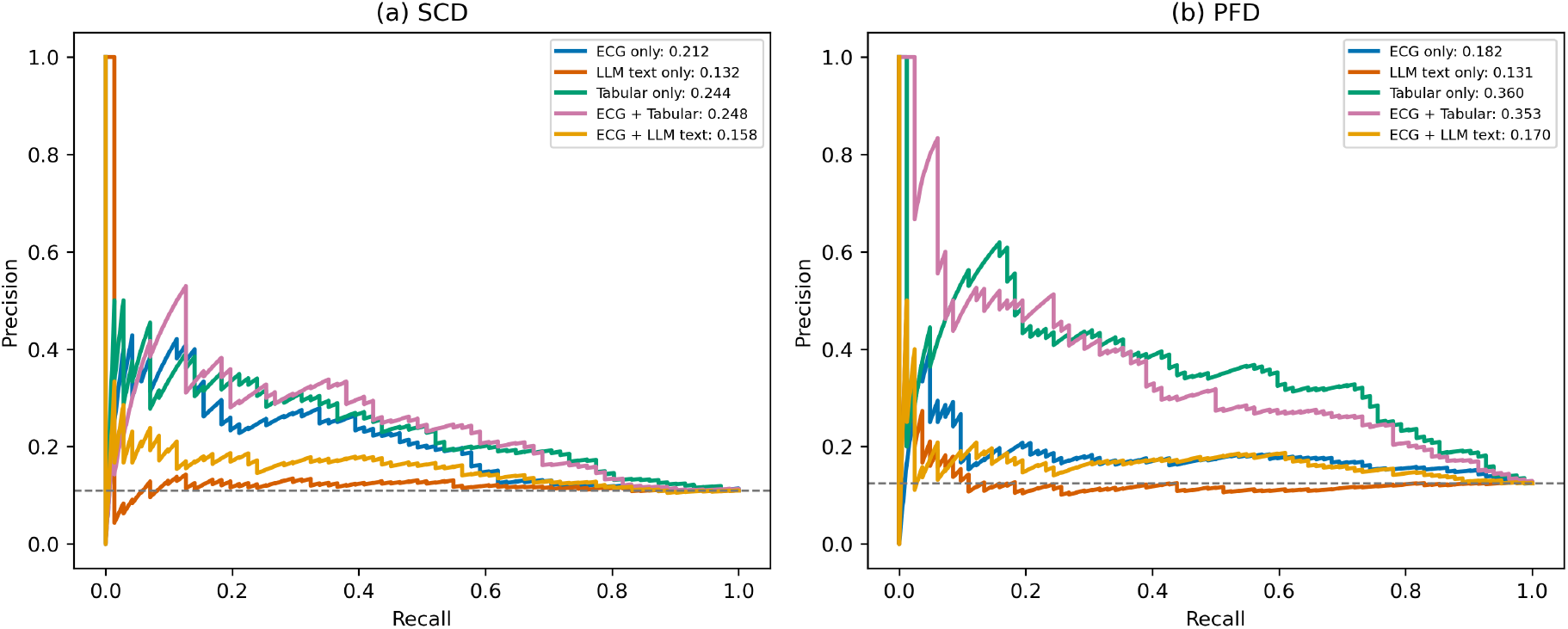
Comparison of PR-AUC curves for unimodal (ECG only, LLM text only, Tabular only) and multimodal (ECG + Tabular, ECG + LLM text) models for sudden cardiac death (a) and pump failure death (b).

**Figure 6:**
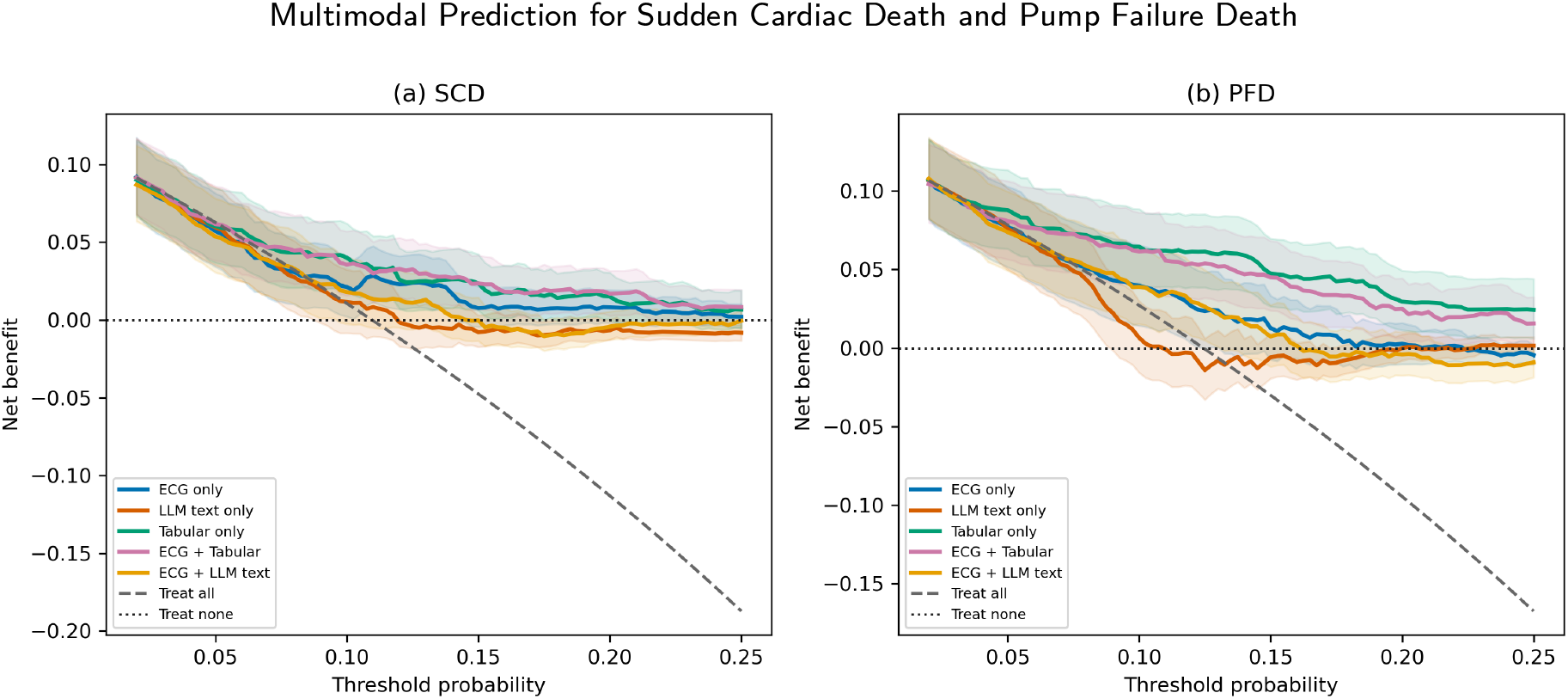
Comparison of DCA curves for unimodal (ECG only, LLM text only, Tabular only) and multimodal (ECG + Tabular, ECG + LLM text) models for sudden cardiac death (a) and pump failure death (b).

**Figure 7:**
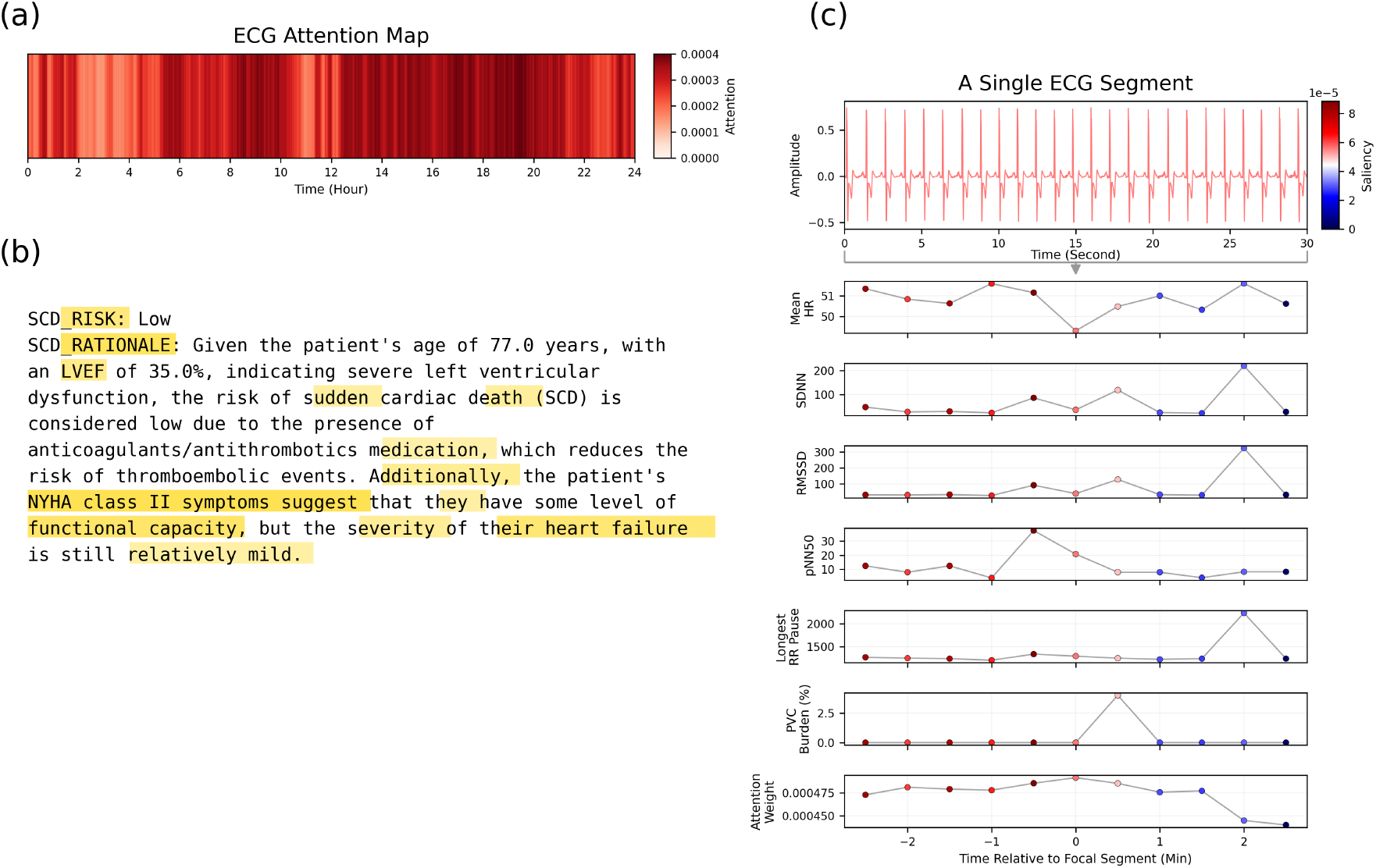
Attention map for the ambulatory ECG (a), gradient-based token-saliency map for the LLM-generated clinical text (b), as well as saliency maps for a select ECG segment and HRV features (c). These maps are for the ECG + LLM text multimodal model in which the patient predicted to experience sudden cardiac death in the next four years.

**Figure 8:**
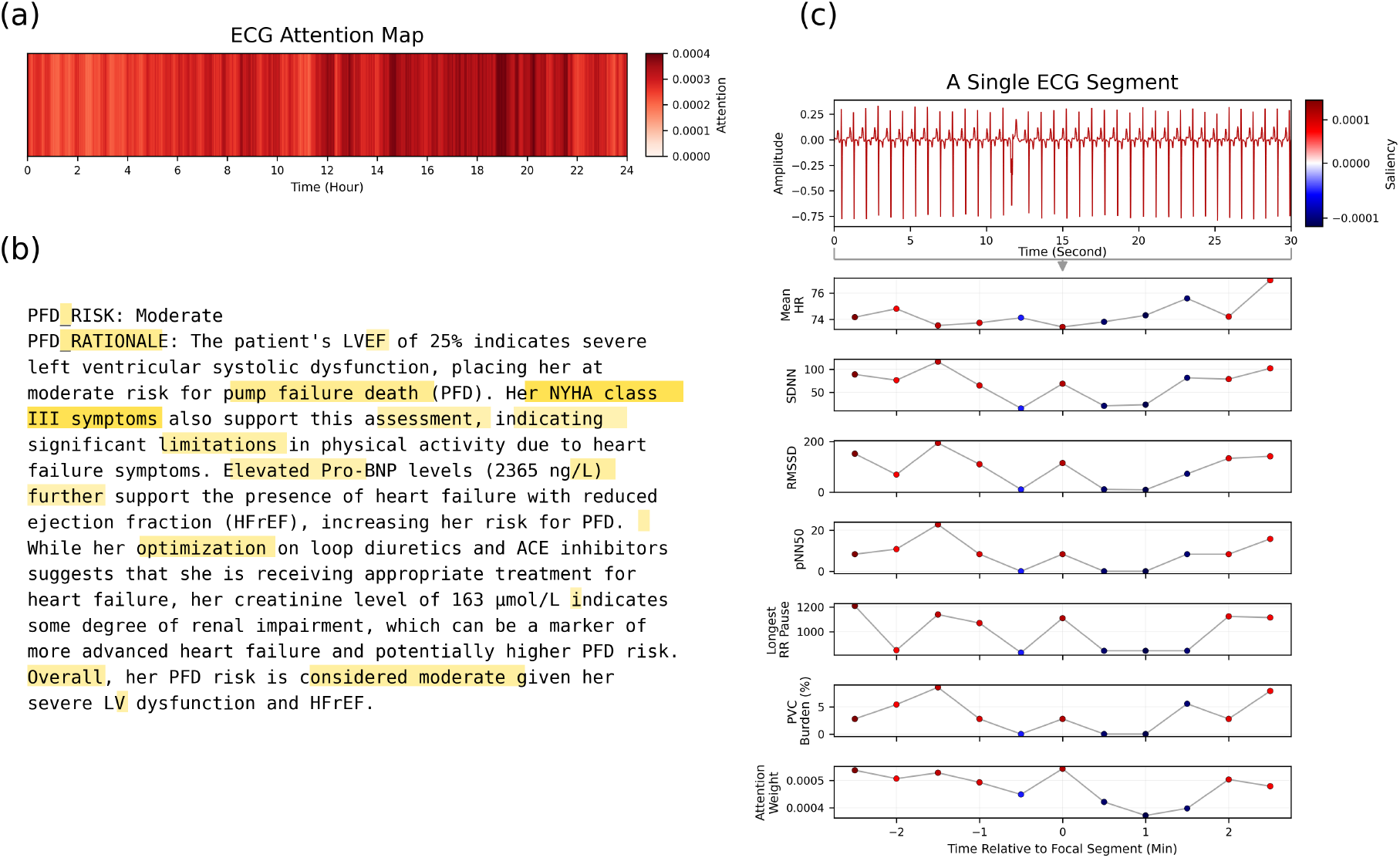
Attention map for the ambulatory ECG (a), gradient-based token-saliency map for the LLM-generated clinical text (b), as well as saliency maps for a select ECG segment and HRV features (c). These maps are for the ECG + LLM text multimodal model in which the patient predicted to experience pump failure death in the next four years.

#### 4.3.1. Unimodal Model Performance

We first evaluated the ECG-derived representation, the full LLM-generated clinical assessment, and the tabular clinical features as separate unimodal inputs. The deterministic text representation was also evaluated as a control for representing the same clinical variables in a fixed format.

For SCD prediction, the tabular model produced the highest point estimates for AUROC and PR-AUC, followed by the deterministic text, ECG-only, and full LLM-text models. Relative to the full LLM representation, the tabular model had a higher AUROC by 0.165 (95% CI, 0.073–0.255; adjusted *p* = 0.052) and a higher PR-AUC by 0.112 (95% CI, 0.041–0.204; adjusted *p* = 0.032). The corresponding AUROC difference did not remain statistically significant after multiplicity correction, whereas the PR-AUC difference did. No other SCD unimodal comparison was statistically significant after Holm correction.

For PFD prediction, the tabular model produced the highest AUROC and PR-AUC, followed by the deterministic text representation. Relative to the full LLM representation, the tabular model had a higher AUROC by 0.321 (95% CI, 0.236–0.404; adjusted *p* = 0.014) and a higher PR-AUC by 0.229 (95% CI, 0.136–0.331; adjusted *p* = 0.014). The tabular model also exceeded the deterministic text representation in AUROC by 0.111 (95% CI, 0.057–0.169; adjusted *p* = 0 014), although their PR-AUC difference was not statistically significant. Deterministic text also outperformed full LLM text for PFD in AUROC, PR-AUC, and Brier score, with adjusted *p* = 0.014 for all three comparisons.

Calibrated Brier scores were generally similar across the SCD models, whereas for PFD the deterministic text representation showed better calibration error than the full LLM representation. Overall, direct tabular and deterministic representations preserved more predictive information than transforming the same clinical variables into a full LLM-generated clinical assessment.

#### 4.3.2. Multimodal Fusion Performance

Next, we evaluated multimodal fusion of ECG representations with text and tabular clinical-data representations. For each modality pairing and outer fold, the LLaMA LLM, biomedical text encoder, downstream text classifier, and fusion architecture were selected using only the corresponding inner cross-validation data. Accordingly, the reported multi-modal performance represents pooled outer-test predictions from the fold-specific selected configurations rather than a single fusion architecture applied across all outer folds. Here, fflLLM textffl refers to the full clinical risk-assessment text generated from structured patient variables by the selected LLaMA LLM and subsequently encoded by the selected biomedical text encoder.

For SCD prediction, ECG + full LLM text achieved an AUROC of 0.591 and PR-AUC of 0.158, with higher point estimates than the unimodal LLM-text model but lower point estimates than the unimodal ECG model. ECG + deterministic text achieved the highest SCD AUROC point estimate overall (0.711), while ECG + tabular clinical features produced the highest PR-AUC point estimate (0.248). None of the prespecified multimodal-versus-corresponding-unimodal comparisons were found to be statistically significant.

For PFD prediction, ECG + full LLM text achieved an AUROC of 0.603 and PR-AUC of 0.170. Its AUROC exceeded that of the unimodal full LLM-text model by 0.136 (95% CI, 0.059–0.215; adjusted *p* = 0.028), although the corresponding PR-AUC and Brier-score differences were not statistically significant. ECG + tabular clinical features achieved an AUROC of 0.756 and PR-AUC of 0.353. Relative to the unimodal ECG model, this corresponded to an AUROC improvement of 0.129 (95% CI, 0.072–0.186; adjusted *p* = 0.014) and a PR-AUC improvement of 0.171 (95% CI, 0.085–0.259; adjusted *p* = 0.041), together with a reduction in Brier score of 0.010 (95% CI, 0.005–0.016; adjusted *p* = 0.041). However, ECG + tabular clinical features did not outperform the unimodal tabular model. ECG + deterministic text performed similarly to deterministic text alone.

Threshold-specific metrics showed a similar outcome-dependent pattern. For SCD, the unimodal ECG model retained the highest accuracy, precision, and specificity, while ECG + deterministic text achieved the highest F1 score among the evaluated models. For PFD, the unimodal tabular model retained the highest sensitivity, NPV, and F1 score, whereas ECG + tabular clinical features achieved the highest accuracy, precision, and specificity. Overall, multimodal fusion provided benefits for selected outcomes and clinical representations rather than uniformly improving performance across all modality pairings.

#### 4.3.3. Exploratory Decision Curve Analysis

Exploratory decision curve analysis evaluated the unimodal ECG, LLM-text, and tabular models and the nested-selected ECG + LLM-text and ECG + tabular models. These models were compared with treat-all and treat-none reference strategies over risk thresholds from 2% to 25%, with pointwise 95% confidence intervals for net benefit obtained from 5,000 patient-level bootstrap replicates.

For SCD prediction, ECG + LLM text did not consistently exceed both reference strategies over the evaluated threshold range. In contrast, ECG + tabular clinical features showed positive net benefit relative to both reference strategies across approximately 8.8%–21.8%. The unimodal tabular model showed favorable net benefit across most of the approximately 8.3%–20.0% range, while the unimodal ECG model exceeded both reference strategies over a narrower interval of approximately 10.8%–14.0%.

For PFD prediction, ECG + LLM text exceeded both reference strategies primarily over a narrow threshold range of approximately 11.0%–13.0%. The unimodal tabular model provided positive net benefit relative to both reference strategies across most thresholds from approximately 6.5% through 25.0%, while ECG + tabular clinical features did so from approximately 6.8% through 24.5%. Thus, the broadest and most sustained regions of positive net benefit were observed for models incorporating the original tabular clinical features.

Because no intervention-specific risk threshold has been clinically validated for these four-year outcomes [52, 53], the 2%–25% interval was selected as an exploratory range encompassing the observed event prevalences and a range of possible risk tolerances. Moreover, the confidence intervals are pointwise and do not provide simultaneous coverage across the entire threshold range. These findings therefore describe comparative model behavior under hypothetical decision thresholds and provide a basis for future external and prospective studies using thresholds linked to specific clinical actions.

#### 4.3.4. Model Attribution and Robustness Analyses

To illustrate the behavior of the ECG + LLM text multimodal models, we present ECG attention and text-saliency visualizations for representative patients predicted to experience SCD and PFD in Fig. 7 and Fig. 8, respectively. Computed attention weights over the full ambulatory ECG (Fig. 7a and Fig. 8a) were distributed throughout the 24-hour recording rather than concentrated within a single isolated interval. These patterns indicate that the temporal aggregation mechanism incorporated information from multiple portions of the recording. For the LLM-generated clinical assessments (Fig. 7b and Fig. 8b), gradient-based token saliency was distributed across multiple portions of the generated text, including clinically relevant descriptors such as LVEF, NYHA class, Pro-BNP, risk labels, and statements regarding heart-failure severity or functional status.

To examine local ECG behavior, representative 30-second segments are shown alongside segment-level HRV features and their corresponding saliency values (Fig. 7c and Fig. 8c). The visualizations illustrate how attention assigned during temporal aggregation can be examined together with changes in heart rate, SDNN, RMSSD, pNN50, longest RR pause, and PVC burden surrounding a focal segment. Taken together, these visualizations provide a complementary way to examine which temporal and clinical features the model emphasizes, potentially helping relate model predictions to recognizable aspects of SCD- and PFD-associated pathophysiology.

We further evaluated the stability of the text-attribution rankings under several controlled perturbations (Fig. 9a,b). Attribution rankings were highly stable to changes in the number of integration steps and remained relatively stable under meaning-preserving input perturbations, including whitespace, punctuation, line-order, and neutral-prefix changes. Stability was lower across repeated outer-training refits with different random seeds, with mean Spearman correlations of 0.596 for SCD and 0.594 for PFD. Randomization of the classifier head produced still lower attribution agreement, with mean Spearman correlations of 0.492 and 0.484 for SCD and PFD, respectively. Overall, similar patterns were observed for both outcomes, indicating that attribution rankings were robust to superficial input changes but more sensitive to variation in fitted model parameters.

**Figure 9:**
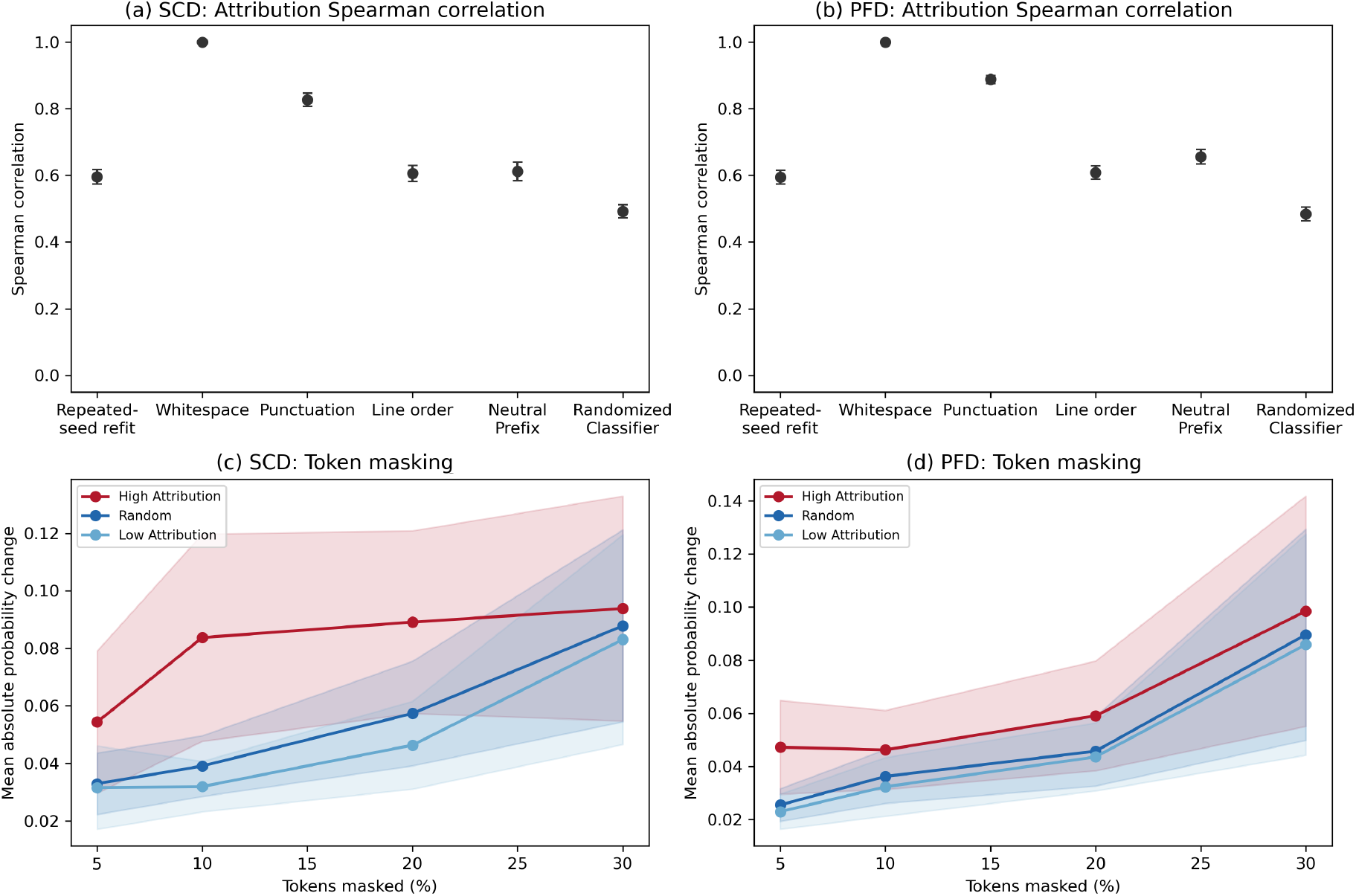
Attribution robustness (a,b) and token-masking analyses (c,d) for SCD and PFD prediction.

Finally, token masking was used to assess whether tokens receiving higher saliency had a greater influence on predicted probabilities (Fig. 9c,d). Among attended, non-special tokens, high- and low-saliency tokens were defined as those ranked at the top and bottom, respectively, by gradient-based token saliency, while random masking sampled an equal number of eligible tokens uniformly without replacement. Masking high-attribution tokens generally produced larger absolute changes in predicted probability than random masking, although the magnitude of this difference varied with the masking fraction. For SCD, the mean high-attribution-minus-random difference was positive at masking fractions of 5%, 10%, and 20%, with 95% confidence intervals excluding zero, but was smaller and not distinguishable from zero at 30%. For PFD, the difference was positive at 5% and 30%, whereas the confidence intervals included zero at 10% and 20%. Together, these analyses provide complementary evidence that the attribution method identifies text regions associated with model output, while also showing that attribution rankings and masking effects are not invariant to changes in the fitted model or perturbation setting.

#### 4.3.5. Representation and Overlap Ablations

We evaluated several representation and ablation conditions to determine whether predictive performance arose from LLM-generated assessment and reasoning, deterministic verbalization of the clinical variables, or direct modeling of the original clinical data. The text conditions included the full LLM-generated risk assessment, risk labels alone, rationales alone, a neutral clinical summary (LLM summary of provided clinical information without assessing SCD or PFD risk), and a deterministic non-LLM template. As a direct clinical-data control, the original numeric and categorical variables were modeled using penalized logistic regression, both without and with the five Holter-derived ECG impressions. Results are summarized in Table 5.

For SCD prediction, the full LLM assessment achieved an AUROC of 0.531 and a PR-AUC of 0.130. The risk-label-only and rationale-only representations did not improve discrimination, while the neutral clinical summary produced modestly higher point estimates, with an AUROC of 0.574 and a PR-AUC of 0.141. In contrast, the deterministic template achieved an AUROC of 0.681 and a PR-AUC of 0.204, approaching the performance of the direct tabular clinical-variable models. The lowest SCD Brier-score point estimate was observed for penalized logistic regression using the clinical variables with ECG impressions.

For PFD prediction, the full LLM assessment achieved an AUROC of 0.459 and a PR-AUC of 0.134. The risk-label-only and rationale-only representations showed similarly limited discrimination, whereas the neutral clinical summary performed better, with an AUROC of 0.620 and a PR-AUC of 0.188. The deterministic template improved further, reaching an AUROC of 0.680 and a PR-AUC of 0.300, although the direct tabular clinical-variable models remained strongest overall.

To evaluate potential overlap with the ECG modality, the five Holter-derived ECG impressions were added separately to the LLM prompts and tabular predictors. For SCD, adding these impressions to the full LLM assessment did not improve discrimination, with AUROC decreasing from 0.531 to 0.516 and PR-AUC from 0.130 to 0.117. In contrast, for PFD, adding ECG impressions increased AUROC from 0.459 to 0.655 and PR-AUC from 0.134 to 0.245. The corresponding tabular AUROCs increased only modestly, from 0.696 to 0.710 for SCD and from 0.787 to 0.792 for PFD. Overall, the ECG-impression fields appeared to contribute substantially more to the LLM representation for PFD than for SCD.

Patient-text correspondence was evaluated by randomly permuting the text representations among patients within each outer-test fold while leaving the ECG representations and outcomes unchanged. The trained fusion models were applied without refitting. This procedure preserved the overall distribution of text embeddings while disrupting the correspondence between each patient and that patient’s text representation.

As shown in Table 6, disrupting patient-text correspondence produced little change in performance. For SCD, correctly paired representations had slightly lower AUROC and PR-AUC than shuffled pairings, with differences of −0.011 and −0.008, respectively, while the Brier-score difference was minimal. For PFD, the corresponding AUROC, PR-AUC, and Brier-score differences were also negligible. None of the permutation tests was statistically significant. These findings suggest that the ECG–LLM fusion model did not derive a measurable performance benefit from preserving patient-specific correspondence in the full LLM-text representation. They also reinforce the broader finding that deterministic and direct tabular representations preserved more usable predictive information than the LLM-generated assessment.

**Table 6.** Patient-text correspondence permutation analysis for ECG + full LLM text. Correctly paired results are pooled outer-test estimates with patient-level bootstrap 95% confidence intervals (CI). Shuffled results are means with 95% permutation intervals. Differences are correctly paired estimates minus shuffled means.

| Outcome | Metric | Correctly paired estimate (95% bootstrap CI) | Shuffled mean (95% permutation interval) | Difference | Permutation p-value |
| --- | --- | --- | --- | --- | --- |
| SCD | AUROC | 0.591 (0.512–0.664) | 0.603 (0.563–0.645) | -0.011 | 0.702 |
| SCD | PR-AUC | 0.158 (0.116–0.231) | 0.166 (0.137–0.208) | -0.008 | 0.635 |
| SCD | Brier score | 0.098 (0.081–0.118) | 0.098 (0.095–0.100) | 0.001 | 0.754 |
| PFD | AUROC | 0.603 (0.546–0.663) | 0.602 (0.571–0.636) | 0.001 | 0.473 |
| PFD | PR-AUC | 0.170 (0.133–0.245) | 0.177 (0.152–0.217) | -0.006 | 0.592 |
| PFD | Brier score | 0.109 (0.091–0.129) | 0.109 (0.107–0.111) | -0.0001 | 0.457 |

## 5. Discussion

We systematically evaluated multimodal fusion strategies that integrate ambulatory ECG-derived representations with clinical text and tabular representations. Multimodal integration was not uniformly beneficial and depended strongly on how the clinical information was represented. For SCD, ECG + deterministic text achieved the highest AUROC, whereas ECG + tabular clinical features achieved the highest PR-AUC. For PFD, the unimodal tabular model remained strongest overall, while ECG + tabular clinical features substantially improved upon the unimodal ECG model but did not outperform tabular features alone. ECG + full LLM text improved upon the unimodal LLM-text model for both outcomes but did not outperform the unimodal ECG model. Among unimodal clinical representations, deterministic and tabular representations generally provided stronger discrimination than the full LLM-generated assessment, particularly for PFD. These findings suggest that clinical context can complement ambulatory ECG representations, but that its predictive value depends on both the outcome and the representation used.

Beyond predictive performance, we present patient-level visualizations of ECG attention and text saliency within the multimodal models. By linking attended ECG segments to corresponding heart rate variability features and clinical text, these analyses highlight features associated with individual model predictions. Attribution rankings were relatively stable to superficial input perturbations but more sensitive to changes in fitted model parameters. Token-masking analyses further showed that masking the highest-ranked tokens by absolute integrated-gradient attribution generally produced larger changes in predicted probability than masking an equal number of randomly selected eligible tokens, although the magnitude of this difference varied by outcome and masking fraction. These analyses provide complementary insight into model behavior while also demonstrating that attribution rankings and masking effects are not invariant to changes in the fitted model or perturbation setting.

Representation and overlap ablations further showed that deterministic text and direct tabular clinical variables preserved more predictive information than the full LLM-generated assessment. Adding ECG impressions had little effect on tabular performance and was more beneficial to the LLM representation for PFD than for SCD. One possible explanation is that rhythm abnormalities may provide additional context for markers of advanced heart-failure severity and impaired cardiac reserve that are more directly related to progressive pump failure, whereas the heterogeneous arrhythmogenic substrate underlying SCD may be less completely captured by these five categorical Holter impressions. However, this interpretation remains exploratory, particularly because adding the same ECG impressions to the tabular models produced only modest performance changes for either outcome. In addition, shuffling patient-text correspondence produced little change in ECG–LLM fusion performance, suggesting that preserving patient-specific correspondence in the full LLM-text representation did not provide a measurable performance benefit under the present analysis. Together, these findings emphasize the importance of evaluating not only whether modalities are combined, but also how each modality is represented and whether it contributes complementary patient-specific information.

Exploratory decision curve analysis showed a similar representation-dependent pattern. Models incorporating the original tabular clinical features exhibited the broadest regions of positive net benefit relative to treat-all and treat-none strategies, particularly for PFD, whereas ECG + full LLM text showed more limited benefit across the evaluated threshold range. Because these thresholds were exploratory and were not linked to a validated intervention strategy, these findings should be interpreted as comparative assessments of potential clinical utility rather than evidence for a specific treatment threshold.

Several limitations should be considered. First, the study was conducted on a single dataset; external validation is necessary to assess generalizability. Second, the relatively limited number of SCD and PFD events may constrain model robustness, particularly for rare-outcome prediction. Third, the current framework relies on engineered HRV features rather than fully end-to-end raw-signal modeling, which may limit capture of more complex waveform-level information. Additionally, the quality and consistency of language model–generated reasoning may introduce variability in text representations. Future work should evaluate these approaches in independent cohorts, investigate end-to-end raw ECG representations, and explore methods for learning more complementary alignment between time-series and clinical representations.

## 6. Conclusion

We present a multimodal AI framework that integrates ambulatory ECG-derived representations with multiple forms of clinical information for four-year prediction of sudden cardiac death and pump failure death. Multimodal fusion was most beneficial for selected outcome–representation combinations rather than uniformly across modalities. For SCD, ECG + deterministic text achieved the highest AU-ROC, while ECG + tabular clinical features achieved the highest PR-AUC; for PFD, the unimodal tabular model remained strongest overall. ECG + full LLM text improved upon LLM text alone but did not outperform the unimodal ECG model. Deterministic and tabular representations generally preserved more predictive information than the full LLM-generated assessment, and patient-text permutation suggested limited measurable benefit from preserving patient-specific correspondence within the full LLM-text representation. Attention, attribution, and token-masking analyses provided complementary insight into model behavior while highlighting the sensitivity of explanations to model and perturbation choices. Overall, these findings emphasize the importance of both modality selection and clinical-information representation in multimodal cardiac outcome prediction and support further evaluation of representation-aware multimodal approaches in external cohorts.

## Data Availability

All data in this manuscript is found at https://physionet.org/content/music-sudden-cardiac-death/1.0.1/.

## CRediT authorship contribution statement

**Steven Swee:** Writing – Original Draft, Validation, Methodology, Software, Investigation, Formal analysis, Data curation, Conceptualization. **Irsyad Adam:** Writing – review & editing, Methodology, Conceptualization, Software, Formal analysis. **Erika Yilin Zheng:** Writing – review & editing, Methodology. **Ethan Ji:** Writing – review & editing, Methodology. **Ding Wang:** Writing – review & editing, Visualization. **William Speier:** Writing – review & editing, Methodology. **Jeffrey Hsu:** Writing – review & editing. **Kai-Wei Chang:** Writing – review & editing, Resources. **Kalyanam Shivkumar:** Writing – review & editing. **Peipei Ping:** Writing - review & editing, Supervision, Conceptualization, Resources, Project administration, Funding acquisition, Formal analysis.

## Declaration of Generative AI and AI-assisted Technologies in the Manuscript Preparation Process

During the preparation of this work, the authors used ChatGPT (GPT-5.6) to review grammar and improve readability. After using this tool, the authors reviewed and edited the content as needed and take full responsibility for the content of the published article.

## Data Availability

Data used in this paper are available on the MIT Physionet database at: https://physionet.org/content/music-sudden-cardiac-death/1.0.1/. The code repository for our study, include LLM responses, is available at: https://github.com/stswee/multimodal_aecg_clinicalfeatures.

## A. Computed ECG Segment Features

Each preprocessed ambulatory ECG was divided into non-overlapping 30-second segments. For each segment, we calculated features describing heart rate variability, beat and rhythm characteristics, heart rate, and signal quality. Table A.1 lists the 28 features supplied to the MIL–TCN model.

**Table A.1:** Segment-level ECG-derived features used by the MIL–TCN model.

| Category | Features |
| --- | --- |
| Time-domain HRV | Mean NN, median NN, SDNN, RMSSD, SDSD, CVNN, CVSD, IQRNN, MadNN, pNN20, pNN50, minimum NN, maximum NN |
| Nonlinear HRV | SD1, SD2, SD1/SD2 ratio, Poincaré ellipse area, sample entropy |
| Frequency-domain HRV | High-frequency power, log-transformed high-frequency power, total power |
| Beat and rhythm | Number of R-peaks, PVC burden, longest RR pause |
| Heart rate | Mean heart rate, heart-rate standard deviation |
| Signal quality | Mean ECG quality, fraction of low-quality signal |

## B. Prompt Templates and Language Model Configurations

Three prompt conditions were evaluated: (1) a full four-year risk assessment without ECG impressions, (2) a neutral clinical summary without ECG impressions, and (3) a full four-year risk assessment with ECG impressions. Each complete prompt was submitted directly to the LLM as a single user message. The common instructions and patient-information template are shown below, and the condition-specific instructions were appended to the common instructions.

**Figure B.1:**
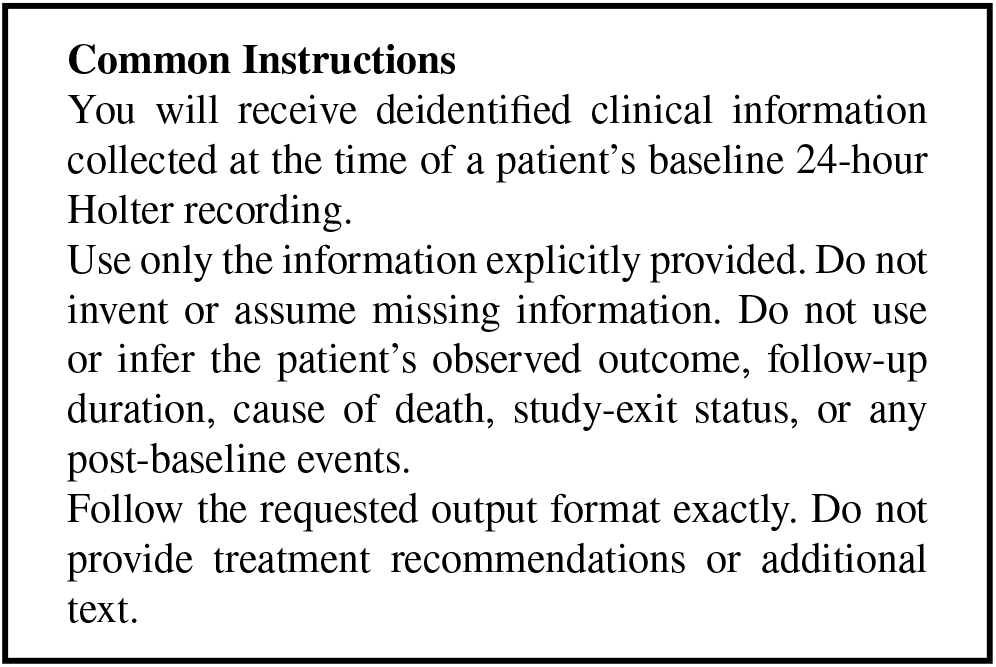
Common instructions used in all LLM prompt conditions.

**Figure B.2:**
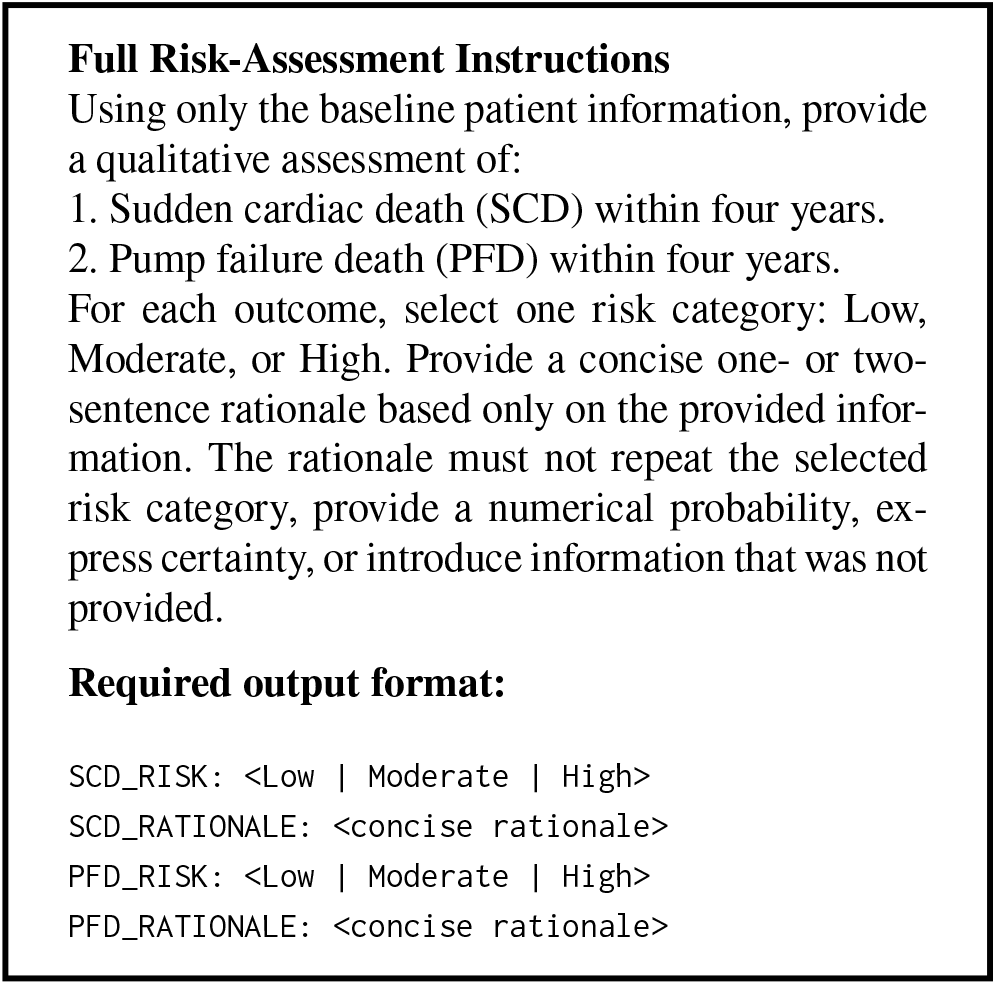
Instructions and output format for the full four-year risk-assessment conditions.

**Figure B.3:**
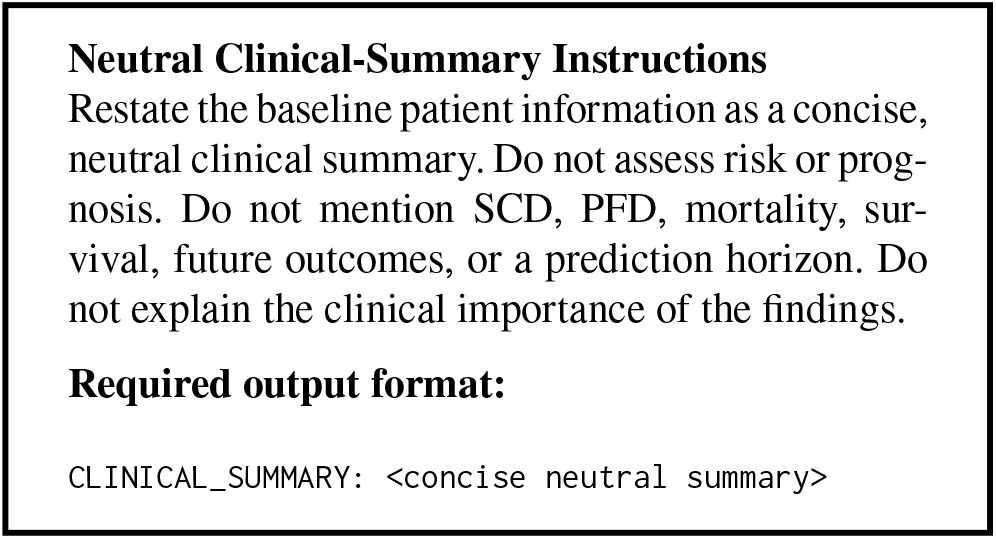
Instructions and output format for the neutral clinical-summary condition.

Patient-specific baseline values were inserted programmatically into the following template. Coded variables were converted into readable clinical descriptions, and unavailable variables were not inferred.

**Figure B.4:**
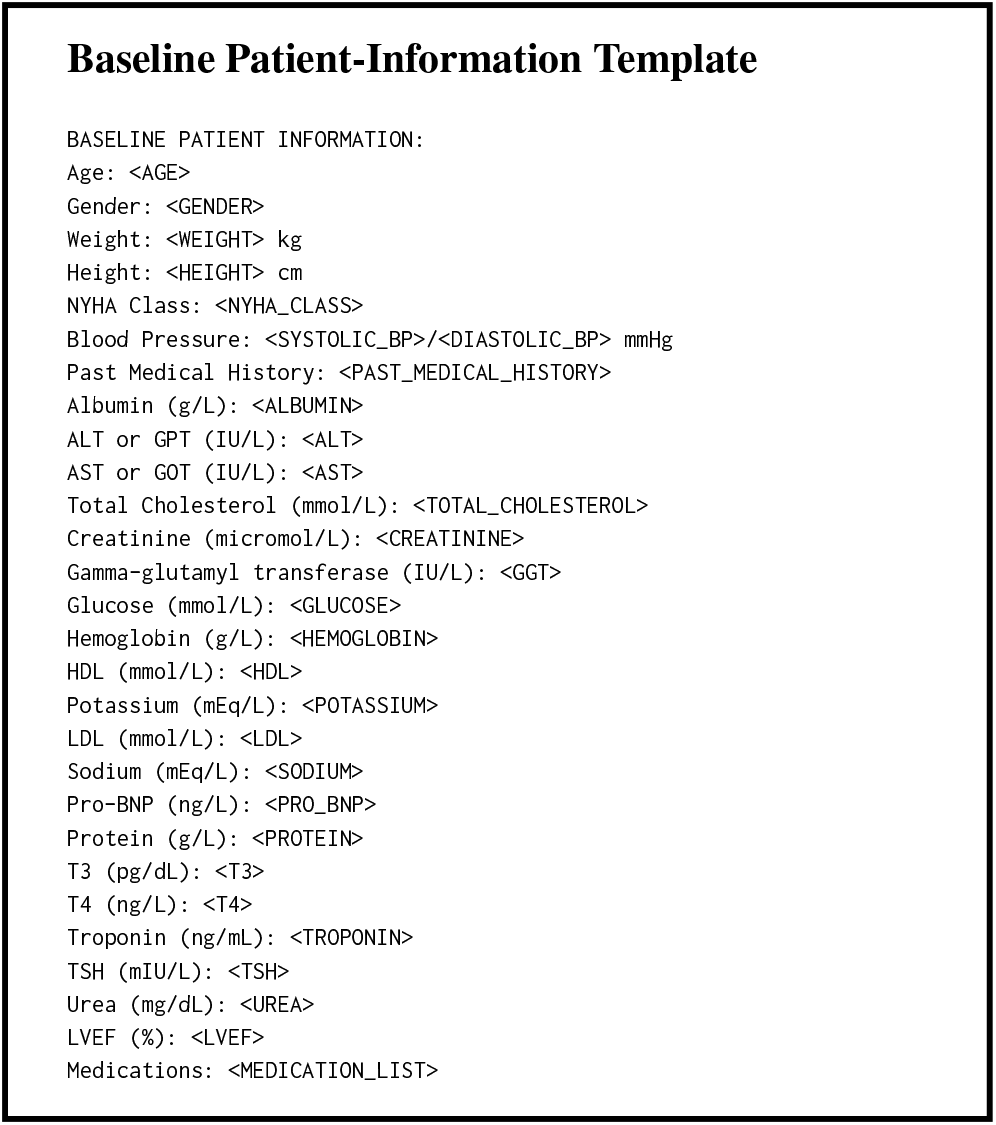
Template used to insert baseline structured clinical variables into each prompt. Angle-bracketed fields indicate patient-specific values inserted programmatically.

The following addendum was included only in the full risk-assessment condition with ECG impressions.

**Figure B.5:**
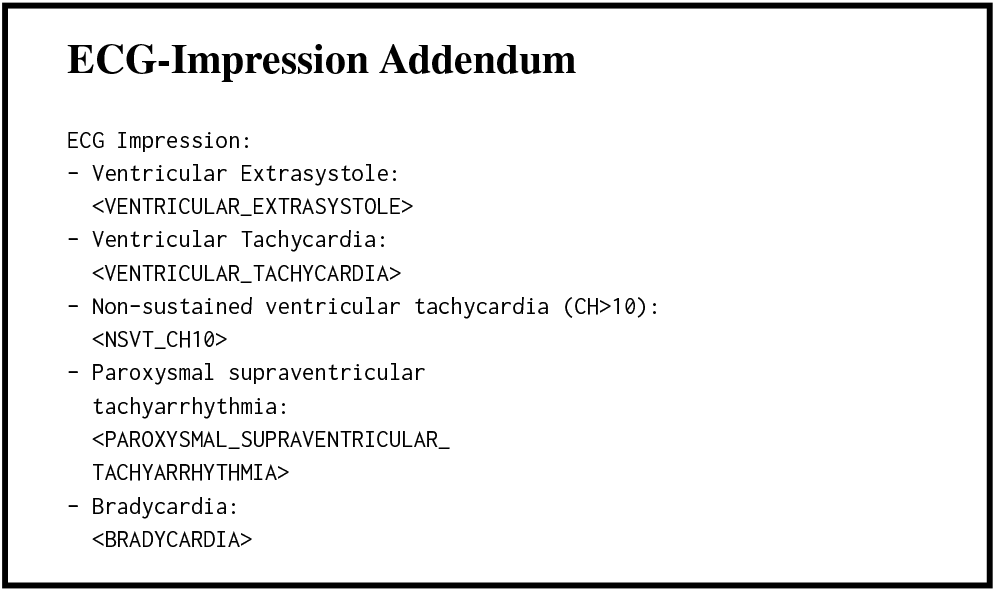
Holter-derived ECG-impression fields included in the full risk-assessment condition with ECG impressions.

The full-risk condition without ECG impressions combined the common instructions, full risk-assessment instructions, and baseline patient-information template. The neutral-summary condition combined the common instructions, neutral-summary instructions, and the same baseline patient-information template. The full-risk condition with ECG impressions additionally included the five Holter-derived ECG impressions.

A deterministic non-LLM representation was constructed directly from the populated baseline patient-information template without submitting it to either LLaMA model. It retained the same structured clinical variables and fixed field order but contained no generated summary, risk classification, or rationale. This representation served as a control for determining whether predictive value arose from converting the structured variables into a fixed textual format rather than from LLM-generated assessment or reasoning.

Risk-label-only representations were constructed from the parsed SCD_RISK and PFD_RISK fields. Rationale-only representations were constructed from the corresponding SCD_RATIONALE and PFD_RATIONALE fields. These representations were derived from the full-risk responses and did not require separate LLM prompts.

Table B.1 reports the exact model snapshots and inference settings used to generate and encode the clinical text. The Hugging Face revision hashes identify the specific model-weight snapshots.

**Table B.1:**
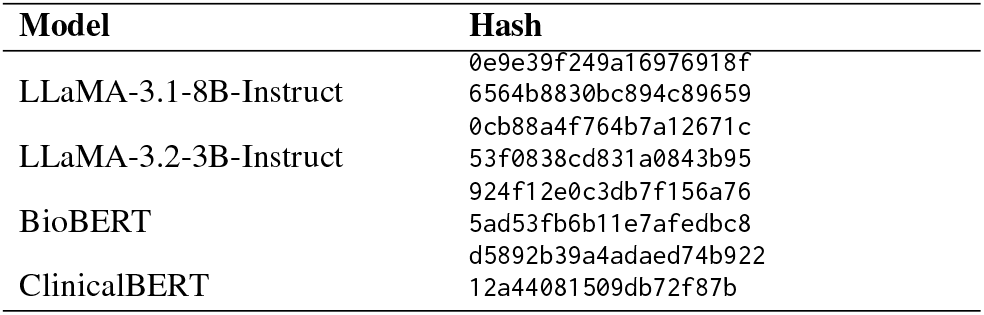
Model snapshots used in the analysis.

## C. Variable Provenance and Modality Overlap

Table C.1 summarizes the categories of clinical information used across the text, tabular, and ECG-impression analysis conditions and identifies potential information overlap with the ECG-derived representation. Variable categories are defined in Table 1.

**Table C.1:**
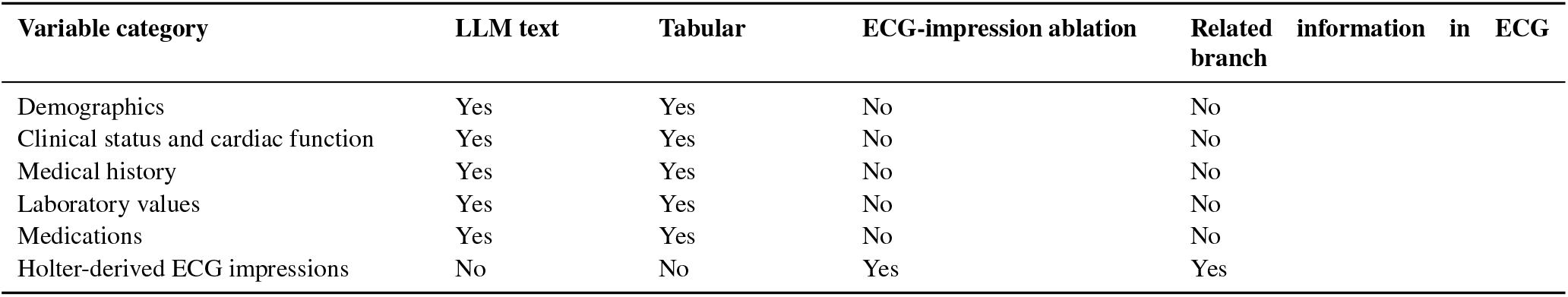
Variable provenance across clinical representations and ECG analyses.

| Variable category | LLM text | Tabular | ECG-impression ablation | Related information in ECG branch |
| --- | --- | --- | --- | --- |
| Demographics | Yes | Yes | No | No |
| Clinical status and cardiac function | Yes | Yes | No | No |
| Medical history | Yes | Yes | No | No |
| Laboratory values | Yes | Yes | No | No |
| Medications | Yes | Yes | No | No |
| Holter-derived ECG impressions | No | No | Yes | Yes |

## D. Hyperparameter Selection

We present the hyperparameters tested in Table D.1, Table D.2, Table D.3, and Table D.4.

**Table D.1:** Hyperparameters tested for the ECG-only (MIL– TCN) classifier.

| Hyperparameters | Values Tested |
| --- | --- |
| Feature Embedding Dimension | 128, 256 |
| TCN Hidden Dimension | 128, 256 |
| TCN Number of Hidden Layers | 3, 4 |
| Encoder Dropout | 0.0, 0.2 |
| TCN Dropout | 0.1, 0.3 |
| Branch Dropout | 0.0, 0.2 |
| Learning Rate | 3e-5, 1e-4, 3e-4, 1e-3 |
| Weight Decay | 0, 1e-5 |

**Table D.2:** Configurations tested for the text embedding classifier.

| Hyperparameters | Values Tested |
| --- | --- |
| LLM Source Model | LLaMA3.1-8B, LLaMA3.2-3B |
| Biomedical Text Encoder | BioBERT, ClinicalBERT |
| Classifier | Linear, MLP |
| MLP Number of Hidden Layers | 1, 2 |
| MLP Hidden Layer Dimension | 128, 256, 512 |
| MLP Dropout | 0.1, 0.2, 0.5 |
| Learning Rate | 1e-4, 3e-4, 1e-3 |
| Weight Decay | 0, 1e-5, 1e-4 |

**Table D.3:** Hyperparameters tested for the conventional tabular classifiers.

| Hyperparameters | Values Tested |
| --- | --- |
| Classifier | Logistic, gradient boosting |
| Logistic Regression Penalty | L2 |
| Inverse Regularization Strength | 0.001, 0.01, 0.1, 1, 10, 100 |
| Gradient Boosting Learning Rate | 0.03, 0.1 |
| Maximum Leaf Nodes | 7, 15 |
| Minimum Samples per Leaf | 20 |
| L2 Regularization | 0, 1 |
| Maximum Boosting Iterations | 300 |

**Table D.4:** Hyperparameters tested for the multimodal classifiers (DC = Direct Concatenation, PC = Projected Concatenation, SG = Scalar Gating, VG = Vector Gating, WS = Weighted Sum).

| Hyperparameters | Values Tested |
| --- | --- |
| Fusion Method | DC, PC, SG, VG, WS |
| Projection Dimension | 128, 256 |
| MLP Number of Hidden Layers | 1, 2 |
| MLP Hidden Dimension | 128, 256 |
| Dropout | 0.2, 0.3 |
| Learning Rate | 3e-4, 1e-3 |
| Weight Decay | 1e-5 |

## E. Cohort Selection

Table E.1 documents the MUSIC cohort selection for our analysis.

**Table E.1.**
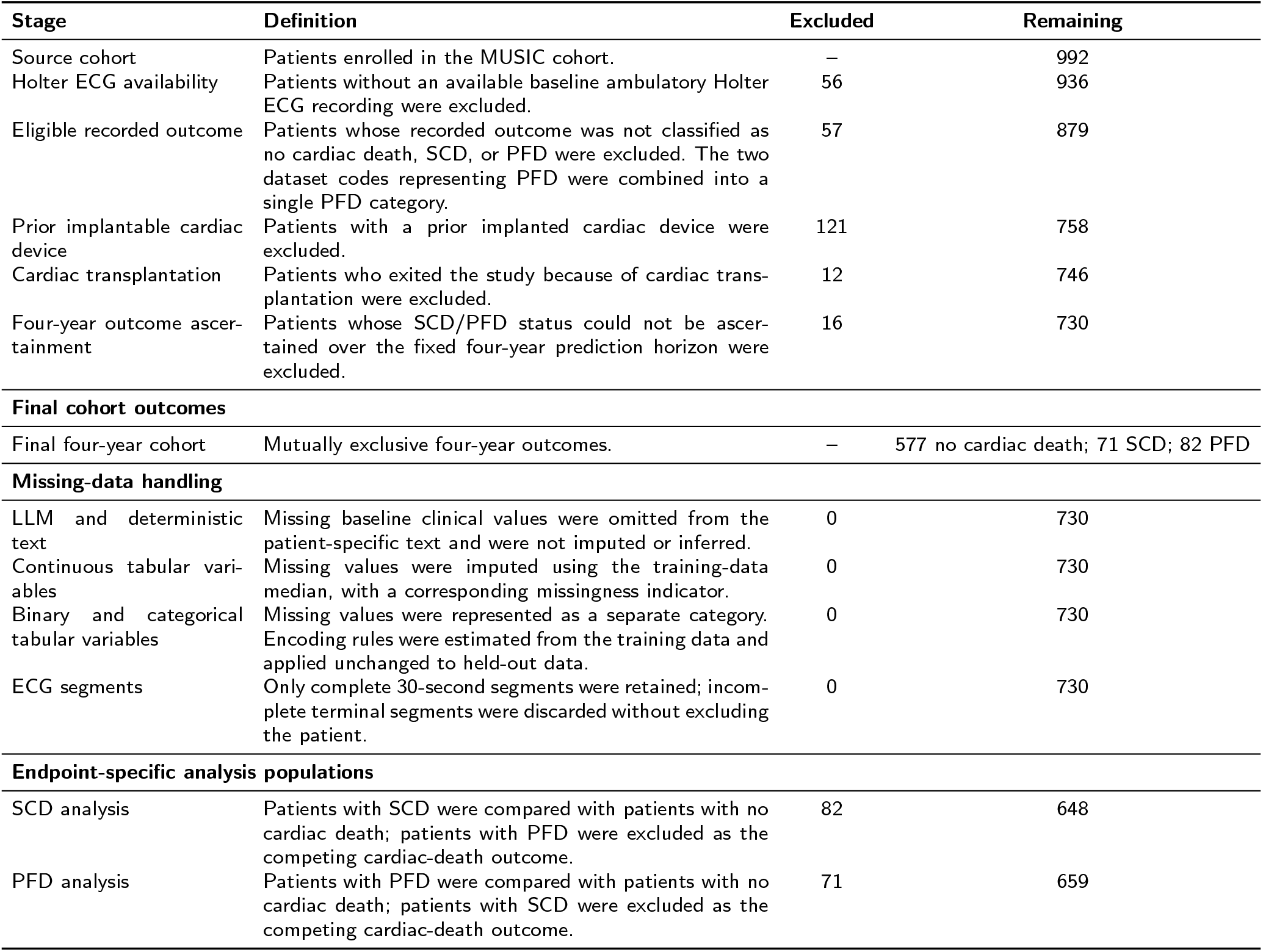
Cohort selection, missing-data handling, and final analysis populations. The baseline 24-hour Holter recording served as the index time, and outcomes were defined over a fixed four-year horizon.

## F. Calibration and Detailed Model Comparison

We present the calibration plots and tabulated calibration values in Figure F.1 and Table F.1, respectively. Paired comparisons between the nested-selected multimodal models and their corresponding unimodal components are reported in Table F.2, and performance across fixed fusion architectures and the nested-selection procedure is summarized in Table F.3. Because nested selection may choose a different fusion architecture in each outer fold, the pooled nested-selected result represents a fold-specific combination of models rather than a single fixed architecture. Consequently, its pooled AUROC can exceed that of every individual fixed architecture, as observed for some SCD modality pairings.

**Figure F.1:**
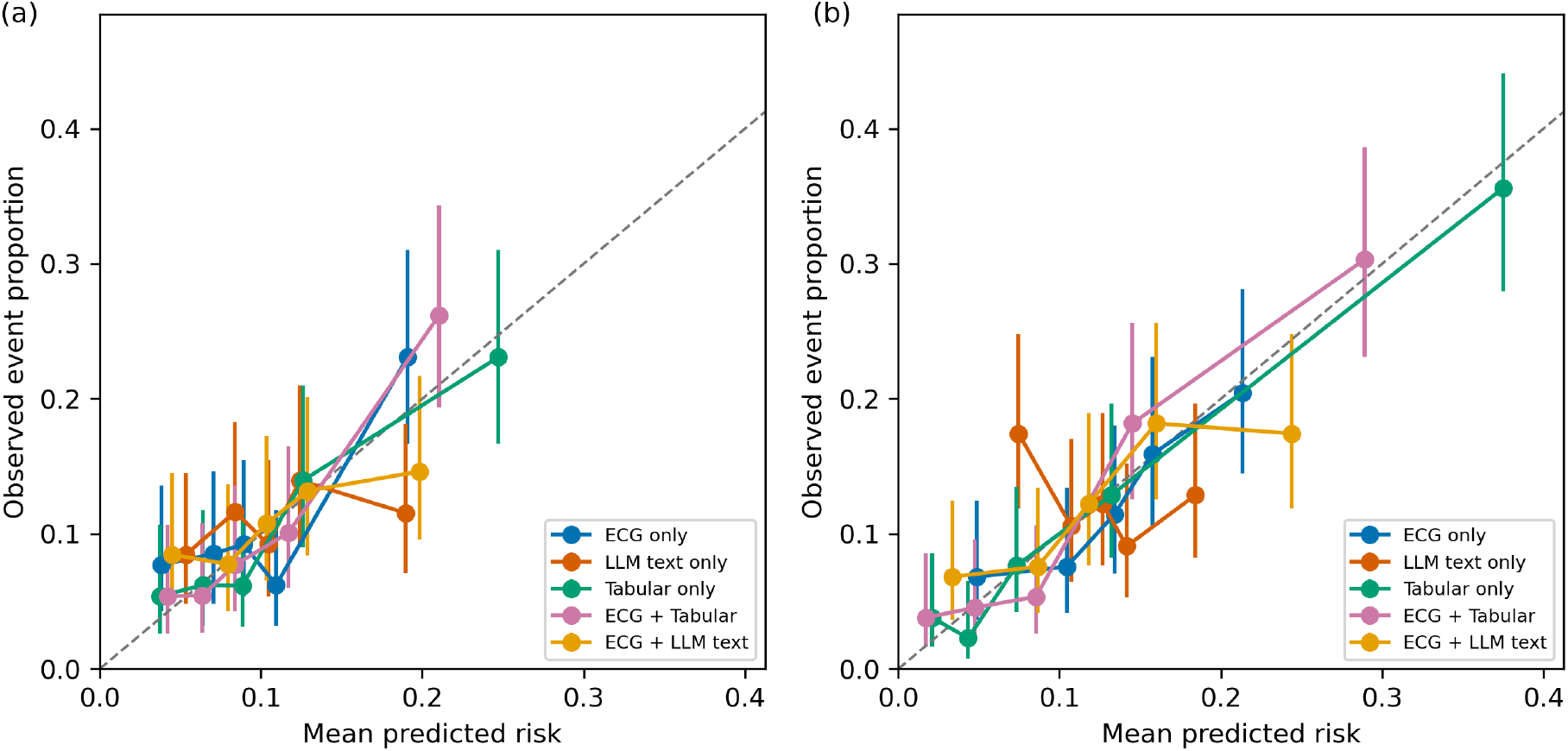
Calibration plots for unimodal and nested-selected multimodal models for four-year sudden cardiac death (a) and pump failure death (b) prediction.

**Table F.1.** Calibration intercept and slope for unimodal and nested-selected multimodal models.

| Model | SCD |  | PFD |  |
| --- | --- | --- | --- | --- |
|  | Calibration intercept | Calibration slope | Calibration intercept | Calibration slope |
| **Unimodal** |  |  |  |  |
| ECG | -0.836<br>(-1.714–0.299) | 0.554<br>(0.188–1.084) | -0.529<br>(-1.377–0.512) | 0.743<br>(0.317–1.312) |
| Full LLM text | -1.877<br>(-2.970–-0.879) | 0.100<br>(-0.373–0.554) | -2.520<br>(-3.865–-1.235) | -0.285<br>(-0.915–0.354) |
| Deterministic text | -0.070<br>(-0.915–0.900) | 0.969<br>(0.580–1.460) | -0.104<br>(-1.041–0.782) | 0.956<br>(0.480–1.473) |
| Tabular clinical features | -0.406<br>(-1.077–0.364) | 0.805<br>(0.501–1.188) | -0.378<br>(-0.812–0.104) | 0.797<br>(0.605–1.041) |
| **Nested-selected multimodal** |  |  |  |  |
| ECG + full LLM text | -1.509<br>(-2.557–-0.454) | 0.269<br>(-0.176–0.772) | -0.889<br>(-1.513–-0.243) | 0.529<br>(0.238–0.869) |
| ECG + deterministic text | -0.044<br>(-0.805–0.695) | 0.921<br>(0.591–1.288) | -0.251<br>(-0.899–0.410) | 0.827<br>(0.516–1.179) |
| ECG + tabular clinical features | 0.283<br>(-0.622–1.182) | 1.108<br>(0.695–1.573) | -0.003<br>(-0.545–0.575) | 0.951<br>(0.681–1.280) |

**Table F.2.** Paired performance comparisons among unimodal and nested-selected multimodal models. Differences are first model minus second model, and *p*-values were adjusted using Holm’s procedure. Bold indicates adjusted *p* < 0.05.

| Comparison | ΔAUROC (95% CI) | $p_{\text{adj}}$ | ΔPR-AUC (95% CI) | $p_{\text{adj}}$ | ΔBrier (95% CI) | $p_{\text{adj}}$ |
| --- | --- | --- | --- | --- | --- | --- |
| **SCD: Unimodal comparisons** |  |  |  |  |  |  |
| LLM text vs ECG | -0.098 (-0.203–0.010) | 1.000 | -0.079 (-0.176–-0.003) | 0.813 | 0.003 (-0.002–0.008) | 1.000 |
| Deterministic text vs ECG | 0.022 (-0.078–0.127) | 1.000 | -0.021 (-0.119–0.070) | 1.000 | -0.002 (-0.008–0.003) | 1.000 |
| Tabular vs ECG | 0.067 (-0.028–0.163) | 1.000 | 0.032 (-0.069–0.135) | 1.000 | -0.004 (-0.011–0.004) | 1.000 |
| Deterministic text vs LLM text | 0.119 (0.032–0.205) | 0.190 | 0.058 (-0.009–0.135) | 1.000 | -0.006 (-0.010–-0.001) | 0.211 |
| Tabular vs LLM text | 0.165 (0.073–0.255) | 0.052 | 0.112 (0.041–0.204) | **0.032** | -0.007 (-0.013–-0.001) | 0.672 |
| Tabular vs deterministic text | 0.046 (-0.034–0.125) | 1.000 | 0.054 (-0.018–0.137) | 1.000 | -0.001 (-0.007–0.004) | 1.000 |
| **SCD: Multimodal vs corresponding unimodal components** |  |  |  |  |  |  |
| ECG + LLM text vs ECG | -0.037 (-0.125–0.052) | 1.000 | -0.054 (-0.125–0.003) | 1.000 | 0.002 (-0.003–0.006) | 1.000 |
| ECG + LLM text vs LLM text | 0.061 (-0.007–0.129) | 1.000 | 0.026 (-0.020–0.084) | 1.000 | -0.002 (-0.004–0.001) | 1.000 |
| ECG + deterministic text vs ECG | 0.083 (0.015–0.154) | 0.525 | 0.031 (-0.049–0.114) | 1.000 | -0.005 (-0.011–-0.001) | 0.864 |
| ECG + deterministic text vs deterministic text | 0.061 (-0.015–0.135) | 1.000 | 0.052 (-0.019–0.130) | 1.000 | -0.003 (-0.007–0.001) | 1.000 |
| ECG + tabular vs ECG | 0.061 (0.001–0.123) | 1.000 | 0.036 (-0.041–0.112) | 1.000 | -0.005 (-0.010–-0.001) | 0.781 |
| ECG + tabular vs tabular | -0.007 (-0.077–0.060) | 1.000 | 0.004 (-0.081–0.082) | 1.000 | -0.002 (-0.007–0.003) | 1.000 |
| **PFD: Unimodal comparisons** |  |  |  |  |  |  |
| LLM text vs ECG | -0.161 (-0.255–-0.068) | **0.023** | -0.051 (-0.115–0.012) | 1.000 | 0.004 (0.001–0.008) | 0.554 |
| Deterministic text vs ECG | 0.049 (-0.039–0.136) | 1.000 | 0.108 (0.019–0.206) | 0.378 | -0.005 (-0.011–0.000) | 0.931 |
| Tabular vs ECG | 0.160 (0.083–0.237) | **0.014** | 0.178 (0.084–0.278) | **0.023** | -0.011 (-0.022–-0.001) | 0.722 |
| Deterministic text vs LLM text | 0.210 (0.120–0.299) | **0.014** | 0.159 (0.068–0.254) | **0.014** | -0.010 (-0.015–-0.004) | **0.014** |
| Tabular vs LLM text | 0.321 (0.236–0.404) | **0.014** | 0.229 (0.136–0.331) | **0.014** | -0.015 (-0.027–-0.004) | 0.239 |
| Tabular vs deterministic text | 0.111 (0.057–0.169) | **0.014** | 0.070 (-0.020–0.161) | 1.000 | -0.006 (-0.015–0.003) | 1.000 |
| **PFD: Multimodal vs corresponding unimodal components** |  |  |  |  |  |  |
| ECG + LLM text vs ECG | -0.024 (-0.087–0.034) | 1.000 | -0.011 (-0.068–0.044) | 1.000 | 0.002 (-0.002–0.006) | 1.000 |
| ECG + LLM text vs LLM text | 0.136 (0.059–0.215) | **0.028** | 0.039 (0.002–0.080) | 1.000 | -0.002 (-0.007–0.001) | 1.000 |
| ECG + deterministic text vs ECG | 0.047 (-0.014–0.107) | 1.000 | 0.058 (-0.012–0.130) | 1.000 | -0.003 (-0.007–0.001) | 1.000 |
| ECG + deterministic text vs deterministic text | -0.002 (-0.059–0.054) | 1.000 | -0.050 (-0.120–0.011) | 1.000 | 0.002 (-0.001–0.006) | 1.000 |
| ECG + tabular vs ECG | 0.129 (0.072–0.186) | **0.014** | 0.171 (0.085–0.259) | **0.041** | -0.010 (-0.016–-0.005) | **0.041** |
| ECG + tabular vs tabular | -0.031 (-0.074–0.013) | 1.000 | -0.007 (-0.083–0.064) | 1.000 | 0.001 (-0.007–0.008) | 1.000 |

**Table F.3.**
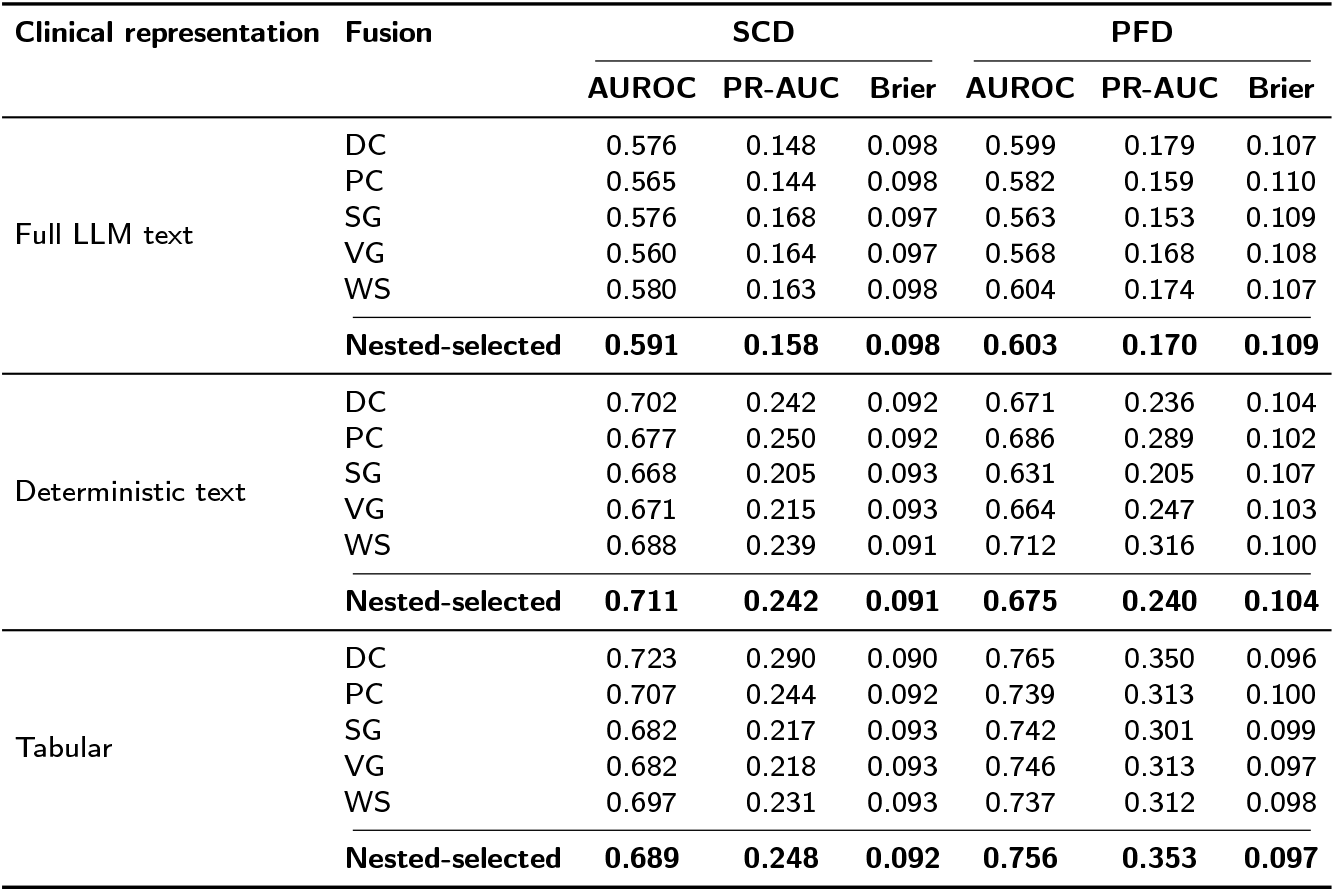
Performance across fixed multimodal fusion architectures and the nested model-selection procedure (DC, direct concatenation; PC, projected concatenation; SG, scalar gating; VG, vector gating; WS, weighted sum).

| Clinical representation | Fusion | SCD |  |  | PFD |  |  |
| --- | --- | --- | --- | --- | --- | --- | --- |
|  |  | AUROC | PR-AUC | Brier | AUROC | PR-AUC | Brier |
| Full LLM text | DC | 0.576 | 0.148 | 0.098 | 0.599 | 0.179 | 0.107 |
|  | PC | 0.565 | 0.144 | 0.098 | 0.582 | 0.159 | 0.110 |
|  | SG | 0.576 | 0.168 | 0.097 | 0.563 | 0.153 | 0.109 |
|  | VG | 0.560 | 0.164 | 0.097 | 0.568 | 0.168 | 0.108 |
|  | WS | 0.580 | 0.163 | 0.098 | 0.604 | 0.174 | 0.107 |
|  | **Nested-selected** | **0.591** | **0.158** | **0.098** | **0.603** | **0.170** | **0.109** |
| Deterministic text | DC | 0.702 | 0.242 | 0.092 | 0.671 | 0.236 | 0.104 |
|  | PC | 0.677 | 0.250 | 0.092 | 0.686 | 0.289 | 0.102 |
|  | SG | 0.668 | 0.205 | 0.093 | 0.631 | 0.205 | 0.107 |
|  | VG | 0.671 | 0.215 | 0.093 | 0.664 | 0.247 | 0.103 |
|  | WS | 0.688 | 0.239 | 0.091 | 0.712 | 0.316 | 0.100 |
|  | **Nested-selected** | **0.711** | **0.242** | **0.091** | **0.675** | **0.240** | **0.104** |
| Tabular | DC | 0.723 | 0.290 | 0.090 | 0.765 | 0.350 | 0.096 |
|  | PC | 0.707 | 0.244 | 0.092 | 0.739 | 0.313 | 0.100 |
|  | SG | 0.682 | 0.217 | 0.093 | 0.742 | 0.301 | 0.099 |
|  | VG | 0.682 | 0.218 | 0.093 | 0.746 | 0.313 | 0.097 |
|  | WS | 0.697 | 0.231 | 0.093 | 0.737 | 0.312 | 0.098 |
|  | **Nested-selected** | **0.689** | **0.248** | **0.092** | **0.756** | **0.353** | **0.097** |

